# Long-term anabolic androgenic steroid use is associated with deviant brain aging

**DOI:** 10.1101/2020.08.19.20177832

**Authors:** Astrid Bjørnebekk, Tobias Kaufmann, Lisa E. Hauger, Sandra Klonteig, Ingunn R. Hullstein, Lars T. Westlye

## Abstract

**Background:** High-dose long-term use of anabolic-androgenic steroids (AAS) may bring a range of health consequences, including brain and cognitive abnormalities. We performed age prediction based on brain scans to test whether prolonged AAS use is associated with accentuated brain aging.

**Methods:** T1-weighted MRI (3D MPRAGE) scans were obtained from male weightlifters with a history of prolonged (n=130) or no (n=99) AAS use. We trained machine learning models on combinations of regional brain volumes, cortical thickness and surface area in an independent training set of 1838 healthy males (18-92 years) and predicted brain age for each participant in our study. Including cross-sectional and longitudinal (mean interval 3.5 years, n=76) MRI data, we used linear mixed effects (LME) models to compare the gap between chronological age and predicted brain age (the brain age gap, BAG) between the two groups, and tested for group differences in the change rate of BAG. We tested for associations between apparent brain aging and AAS use duration, administration pattern and dependence.

**Results:** AAS users had higher BAG compared to weightlifting controls, associated with dependency and longer history of use. Group differences in BAG could not be explained by other substance use, general cognitive abilities or depression. Longitudinal data revealed no evidence of accelerated brain aging in the overall AAS group, though accelerated brain aging was seen with longer AAS exposure.

**Conclusions:** The findings suggest that long-term high dose AAS use may have adverse effects on brain aging, potentially linked to dependency and exaggerated use of AAS.

## Introduction

Anabolic-androgenic steroids (AAS) are a family of hormones that comprise testosterone, and hundreds of synthetic derivatives of testosterone (1). The intake of supraphysiological doses of AAS in combination with strength training increases lean muscle mass and strength (2). These are desired effects for athletes and bodybuilders where widespread use was seen from the 1950s, before it spread to the general population around the 1980s. AAS use brings a range of health and social consequences (3, 4). Yet, long-term effects on brain health and cognition are understudied, which is paradoxical since sex steroids readily pass the blood-brain barrier and affect the central nervous system (CNS).

The biological action of AAS and their metabolites are primarily mediated via the androgen receptors (AR), however many will also exert physiological effects via estrogen receptor pathways, upon aromatization (5, 6). Sex steroid receptors are widely expressed in the brain, and abundantly in regions such as the brainstem, hypothalamus, amygdala, striatum, hippocampus and the cerebral cortex (7-9). High-dose AAS administration typically involves a complex pattern where different testosterone compounds and other AAS are co-administered with doses equivalent to 250-5000 mg/week, which is 5-100 times greater than the natural male production (10). Administration of supraphysiological AAS-doses causes suppression of the hypothalamic-pituitary gonadal axis, reducing the production of endogenous testosterone, luteinizing and follicle-stimulating hormones. The administration periods typically last for several weeks or months, separated by drug-free intervals with the intention to allow the hormonal system to recuperate (11). However, it seems that continuous use persisting for years have become more common (12-16), likely as a way to relieve the abstinence symptoms that often occur upon cessation (17, 18).

Growing evidence suggests that high-dose long-term exposure to AAS have harmful effects on the brain. Cell culture studies have shown neurotoxic effects of various sorts of AAS (19-24), in particular in response to high dosages mimicking those taken by bodybuilders and recreational athletes (23, 24). Contrary, neuroprotective effects of physiological doses of testosterone have been observed (25, 26). Moreover, AAS use frequently causes cardiomyopathy (27, 28), atherosclerotic disease (27), prolonged hypogonadism (upon withdrawal) (29, 30), lower LDL cholesterol level (31), impaired insulin sensitivity (32), and occasionally toxicity to liver and kidney (33), with potential implications for brain health (34, 35).

Emerging evidence from field studies suggests that prolonged high-dose AAS use is associated with aberrant brain aging. For instance brain imaging has revealed that long-term AAS-use is associated with structural, neurochemical (36), and functional brain differences (36-38), including smaller gray matter, cortical and putamen volume, and thinner cerebral cortex (37). Also, compared to non-using weightlifters, AAS-exposed weightlifters have shown to perform poorer on tests assessing working memory (12, 39, 40), executive functions (12, 40, 41), learning and memory (12, 39, 41), processing speed and problem solving (12, 40). Although correlational, such findings have led to the hypothesis that high-dose AAS users are at risk for accelerated brain aging (42, 43).

The effects of AAS use show substantial inter-individual heterogeneity. Some users exhibit little or no psychological or medical symptoms, while others demonstrate multiple mental and medical consequences following long-term use (11, 44). The range and severity of adverse effects may increase with the burden of use (19), and are particularly pronounced in users fulfilling the criteria for AAS-dependence (1, 15, 45). This includes seemingly more pronounced effects on MRI-based measures of cerebral cortical structure (37, 45), self-reported memory problems (12, 41), and impaired executive (40) and memory functions (12, 39) in dependent users. However, group-level differences may disguise substantial individual differences.

Machine learning has offered novel individual-based predictions based on neuroimaging data (46). For example, training a model to find relationships between brain scans and chronological age allows you to predict the age from unseen brain images with high accuracy (47, 48). The difference between the predicted and chronological age, termed the *brain age gap* (BAG), serves as a surrogate marker of individual brain health and individual differences in brain maturation and aging (49, 50). In adults, an older brain age compared to chronological age has been linked with cognitive impairment (51), cardiovascular risk factors (34), mortality (52), dementia (53), and several other common brain disorders, with regionally differing patterns (54). Some evidence suggests that healthy lifestyle factors are associated with a younger looking brain, with negative correlations between BAG and level of education and the daily number of flights of stairs climbed (55). Contrary, drug abuse and addiction has been associated with premature brain aging (56-58) and early onset of age-related disease (59). While recent studies have documented associations between cumulative exposure to sex hormones and brain age in middle-aged and elderly women (60), the effects of long-term exposure of supraphysiological doses of testosterone and AAS on brain aging have not been studied.

In a sample of 130 AAS users and 99 weightlifting controls (WLC), we used cross-sectional (n=229) and longitudinal (n=76) data to test the hypothesis of higher relative brain age and higher rates of brain aging in AAS users compared to WLC. We also tested for associations between brain age and AAS use severity, duration, administration (cycling versus continuous use) and dependence.

## METHODS AND MATERIALS

### Participants

Demographics and clinical characteristics of the sample are summarized in table 1. The sample is part of a longitudinal study investigating effects of long-term AAS use on brain morphology, cognitive functioning, and emotional processing. Data collection was performed during 2013-2015 and 2017-2019. We recruited males engaged in heavy resistance strength training who were either current or previous AAS users reporting at least one year of cumulative AAS exposure (summarizing on-cycle periods) or who had never tried AAS or equivalent doping substances. Participants were recruited through webpages and forums targeting people partaking in heavy weight training, bodybuilding, and online forums (open and closed) directly addressing AAS use. In addition, posters and flyers were distributed at select gyms in Oslo. Prior to enrollment all participants received an information brochure with a complete description of the study. The study was approved by the Regional Committees for Medical and Health Research Ethics South East Norway (REC) (2013/601), all research was carried out in accordance with the Declaration of Helsinki, and written informed consent was collected from all subjects. The participants were compensated with 1000 NOK at time point 1 (TP1) and 500 NOK at time point 2 (TP2).

**Table 1.**
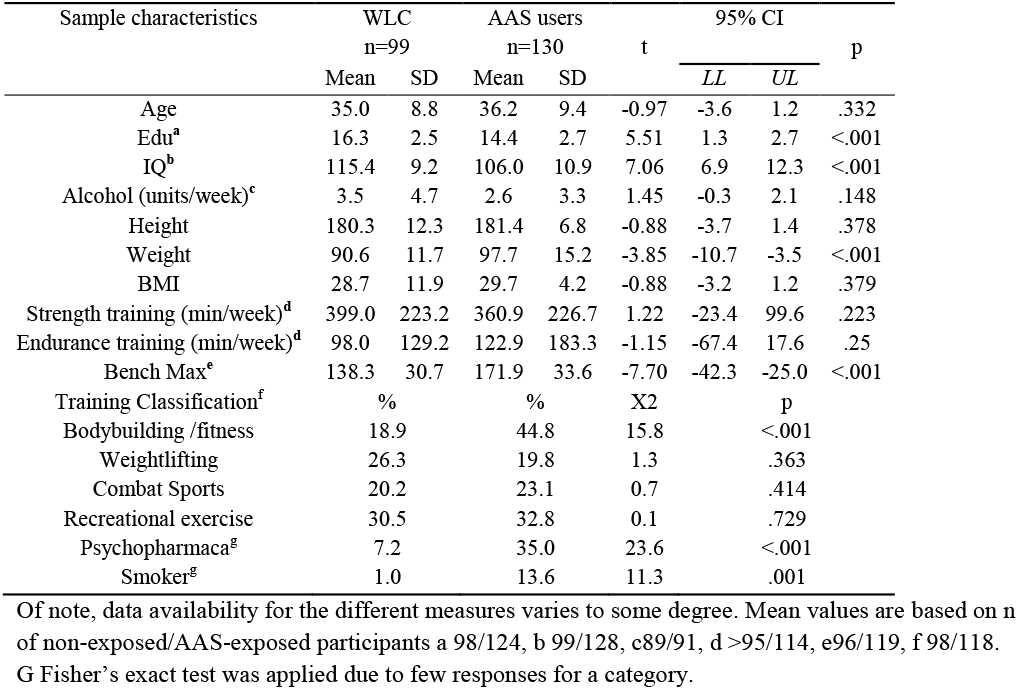
Demographics, Sports Information, Substance Abuse, and Use of Psychopharmaca

In total 139 AAS users and 109 WLC underwent brain MRI. 19 participants were excluded. Among AAS users, two participants did not fulfill the inclusion criteria of having 1 year cumulative exposure, one was excluded because of a previous head injury that had caused coma, one due to poor scan quality, two due to IQ <80, and two due to missing background information. Among WLC one was excluded due to epilepsy, two did not match the AAS group on strength training regimens, and three were excluded due to missing background information. Furthermore, three WLC were excluded due to clinical significant abnormalities based on a neuroradiological examination. In addition one 73 year old AAS user and a 75 year old WLC were excluded due to their substantially higher age than the rest of the sample, which may influence the brain age models and the findings. Therefore, our final sample comprised 130 AAS users and 99 WLC. Among those, 36 AAS users and 40 WLC were scanned at TP2, on average 3.5 years after TP1.

### Image Acquisition

MRI data was collected using a 3.0T Siemens Skyra scanner (MAGNETOM Skyra; Siemens AG, Erlangen, Germany) equipped with a 20-channel head coil. Anatomical 3D T1-weighted magnetization-prepared rapid acquisition gradient-echo (MPRAGE) sequences with the following parameters were used for volumetry and cortical surface analyses: repetition time 2300 ms; echo time 2.98 ms; inversion time 850 ms; flip angle 81°; bandwidth 240 Hz/pixel; field of view 256 mm; voxel size 1.0 * 1.0 * 1.0 mm; 176 sagittal slices; acquisition time 9:50 min. Scan quality was inspected at the scan session and rerun in case of movement.

### MRI processing and brain age estimation

All datasets were processed using Freesurfer v.5.3 (https://surfer.nmr.mgh.harvard.edu/; (61), and segmentations and reconstructions were visually inspected and edited if needed. The training set for brain age estimation included MRI scans from 1838 healthy males from different cohorts (mean age 46 years, (±) sd 20 years, age range 18-92 years) obtained from several publicly available datasets and processed in the same pipeline. The age distributions for the training set and our cohort is shown in Figure 1a, and information about included datasets are shown in Supplementary table 1.

**Figure 1.**
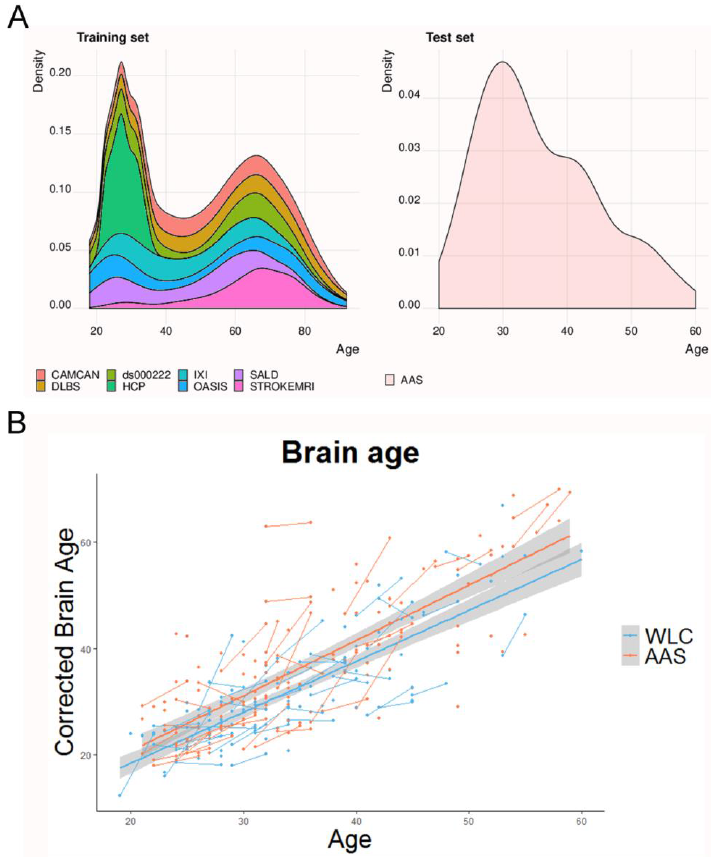
Age distribution and predicted brain age as a function of age A) The age distributions for the training set and our cohort. B) Predicted global brain age corrected for age, as a function of chronological age. The fit lines represent the best linear fit within each group, and the points connected by lines represent individual change in BAG between the two MRI scans for each individual.

A machine learning model was trained to predict brain age based on volume, area and thickness data following a recent implementation (54). The features were derived from the Human Connectome Project cortical parcellation (62), comprising 180 regions of interest per hemisphere for thickness, area, and volume. In addition, we used subcortical and cerebellar volumes from Freesurfer. The full set comprised 1,118 features in total. We used the extreme gradient boosting package xgboost in R to train machine learning models for brain age estimation. In line with recent work, the learning rate was pre-set to eta=0.01 and the optimal number of rounds (nrounds) were determined in a nested cross-validation loop (54).

For all participants, brain age and BAG were estimated using either features from the whole brain or subregions (54, 63), including occipital, frontal, temporal, parietal, cingulate, insula, or cerebellar/subcortical features, based on the lobe parcellation labels from Freesurfer (61). We corrected for a well-known bias in age prediction (64) using a procedure described in (65). Briefly, the association between BAG and age was estimated using linear models including relevant covariates, and the resulting parameter estimate reflecting the linear association between BAG and chronological age was used to adjust the individual brain age estimates prior to recalculation of BAG.

### Interviews and screening instruments

Demographics and clinical data were assessed using self-report questionnaire and a semi-structured interview. Current and previous non-AAS substance use were assessed with Alcohol Use Disorders Identification Test (AUDIT) (66), the Drug Use Disorders Identification Test (DUDIT) (67), and the drug and alcohol dependence scales from the Millon Clinical Multiaxial Inventory-III (MCMI-III), where a composite scores of substance use were computed from the mean score of these z-transformed subtests. The depression scale from the MCMI-III was used to covary for depressive symptoms. “Total lifetime AAS dose” ingested was calculated as the life-time average weekly dose reported and life-time weeks of AAS exposure, in line with previous studies (1, 68, 69). Intelligence Quotient (IQ) was assessed using the Wechsler Abbreviated Scale of Intelligence (WASI) (70).

### Doping analysis

Urine samples were collected and analyzed for AAS and some antiestrogens using gas and liquid chromatography coupled to mass spectrometry at the WADA accredited Norwegian Doping Laboratory at Oslo University Hospital (71). The criteria used to determine AAS use were: 1) urine samples positive for AAS compounds 2) a T/E ratio > 15 equivalent to previous work (37, 71). Other compounds, including stimulants and remaining antiestrogens were analyzed with liquid chromatography and mass spectrometry.

### Statistical Analysis

Group differences in demographic data were evaluated with two-tailed independent sample t-tests and Chi-square and Fisher’s exact tests for categorical data. To assess group differences in global and regional BAG linear mixed effects (LME) models analyses were performed using the *lme* function in the R (72) package lme4 (73). In fitting the model, we entered timepoint (TP) and age as fixed effects. Participant ID was entered as a random effect (intercept). Visual inspection of residual plots did not reveal obvious deviations from homoscedasticity or normality. The significance threshold was set at p < 0.05, corrected for multiple comparisons using false discovery rate (FDR) adjustment (74).

#### Sensitivity analyses

Similar LME models including a group by time interaction was run to test for differences in the rate of change between AAS users and WLC. In addition, to test for confounding effects of cognitive ability, depression and non-AAS substance use, the main analyses were rerun including IQ, depressive scores and a composite score of non-AAS substance use as additional covariates. As we were primarily interested in long-term exposure and since stricter inclusion criteria have previously been applied (29, 32), we rerun the main analyses after including only AAS users with more than two years of AAS use.

Next, similar LME analyses were conducted to test for differences between subgroups of AAS users. 1) Use category: WLC, AAS users fulfilling the criteria for AAS-dependence and non-dependent users, 2) Use pattern: WLC, AAS users practicing a continuous way of administrating AAS versus users administrating AAS in cycles, and 3) Use state: WLC, current and previous AAS users. 4) Use length: AAS users with < 10 years of exposure and users with ≥10 years history of AAS exposure.

Lastly, as only ∼50% of the sample took part at TPII, we conducted linear models controlling for age to examine if BAG at baseline was associated with study dropout.

## Results

### Demographics and user characteristics

Table 1 summarizes key clinical and demographic characteristics. Years of education and IQ were higher among WLC, and AAS users were heavier and stronger than WLC. The use of prescribed psychotropic medication was significantly higher among AAS users, where antidepressants and anxiolytics were the typical preparations prescribed (not shown). The majority of users (65%) and non-users (93%) reported no previous or current use of prescribed psychotropic medication.

### Characteristics of AAS use

The average duration of AAS use at baseline was 10.6 years (SD=7.7), and mean age of onset was 22 years (SD=6.6, range 12-52). Mean weekly AAS dosage was 1023 mg (SD=656, range 100-3750), and mean calculated lifetime dose was 444 g (SD=452 g, range 20-2016 g). Continuous AAS administration was reported by 43 (33.1 %) users, and 78 (60.0%) reported a cycling pattern, rotating between periods “on” and “off” AAS. The remaining 9 (6.9%) were either on testosterone replacement therapy, had missing details regarding administration pattern or was difficult to classify. 77 (59.2%) AAS users fulfilled the criteria for AAS-dependency, and 87 (67%) had used AAS within the past 6 months and defined as current users. Current users had longer history of AAS use although they debuted later compared with past AAS users. No differences in extent of use measures were seen between cyclical versus continuous users, whereas dependent users had used longer, debuted earlier and used higher AAS doses compared to non-dependent users (Table S2).

None of the 99 WLC tested positive for AAS or had T/E ratio above threshold, whereas tests indicative of AAS use were seen in 78.2% (n=68) of current users, and in 7.5 % (n=3) of previous users. The positive tests among previous users could be compatible with use back in time, stated by the participants, and one test with elevated T/E ratio was consistent with reported medical use of TRT. The mean T/E ratio for the groups were 1.4 (SD=1.6, range 0.1 – 10.0) for WLC (n=99), 44.8 (SD=50.6, range 0.1 – 226.0) for current users (n=82), and 2.8 (SD=7.9, range 0.0 – 50.4) for previous users, where previous users and WLC were significantly different from current users (df=220, t=-7.2, p<.001). The frequency of the specific anabolic-androgenic steroids found in the urine sample are summarized in Figure S1 along with a summary of the most popular compounds based on self-reports.

### Brain age prediction

A 10-fold cross validation on age prediction in the training set confirmed high accuracy of the model, with correlations between chronological age and predicted age ranging from *r*=.926 (MAE=5.76, RMSE=7.57,) for the global model to *r*=.76 (MAE=10.05, RMSE=12.94) for the model based on occipital features (Table. S3). Figure 1 shows predicted age plotted as a function of chronological age for the test set of AAS users and WLCs, and Table S3 summarizes the prediction accuracies.

### Associations between AAS use and BAG

Table 2 summarizes the results from the LME models. Significant main effects for group were found for the global BAG (β) (305) = 3.29, t=3.58, p_FDR_<.001), and for the frontal, temporal, insula, cingulate and occipital BAGs. An examination of the fixed effects estimates showed higher BAG in AAS users compared to WLC in all regions. There were no significant main effects of time or age.

**Table 2:** Main model.

|  | Fullbrain | Frontal | Temporal | Insula | Cingulate | Parietal | Occipital | Subcortical |
| --- | --- | --- | --- | --- | --- | --- | --- | --- |
| Group AAS | 3.288***<br>(0.918) | 3.743**<br>(1.177) | 2.573*<br>(1.046) | 2.483*<br>(0.979) | 2.613*<br>(1.185) | 2.033*<br>(1.025) | 2.885*<br>(1.154) | 1.906<br>(1.064) |
|  | 3.580<br><.001 | 3.180<br>0.008 | 2.460<br>0.024 | 2.535<br>0.024 | 2.205<br>0.037 | 1.984<br>0.055 | 2.499<br>0.024 | 1.790<br>0.075 |
| Time | 0.260<br>(0.513) | 0.221<br>(0.720) | -0.846<br>(0.563) | -0.879<br>(0.552) | -0.436<br>(0.685) | 0.008<br>(0.554) | 0.874<br>(0.701) | 0.796<br>(0.522) |
|  | 0.507<br>0.817 | 0.307<br>0.867 | -1.504<br>0.36 | -1.592<br>0.36 | -0.636<br>0.817 | 0.015<br>0.988 | 1.246<br>0.432 | 1.527<br>0.36 |
| Age | -0.021<br>(0.050) | -0.007<br>(0.064) | -0.033<br>(0.056) | -0.020<br>(0.053) | 0.007<br>(0.064) | -0.015<br>(0.055) | -0.025<br>(0.062) | -0.056<br>(0.057) |
|  | -0.415<br>0.916 | -0.105<br>0.916 | -0.590<br>0.916 | -0.385<br>0.916 | 0.113<br>0.916 | -0.280<br>0.916 | -0.394<br>0.916 | -0.975<br>0.916 |
| Observations | 305 | 305 | 305 | 305 | 305 | 305 | 305 | 305 |
| Log Likelihood | -986.417 | -1,070.714 | -1,022.649 | -1,007.022 | -1,067.389 | -1,016.988 | -1,064.043 | -1,019.256 |
| Akaike Inf. Crit. | 1,984.834 | 2,153.427 | 2,057.298 | 2,026.044 | 2,146.777 | 2,045.976 | 2,140.085 | 2,050.512 |
| Bayesian Inf. Crit. | 2,007.155 | 2,175.749 | 2,079.620 | 2,048.366 | 2,169.099 | 2,068.298 | 2,162.407 | 2,072.834 |

Linear mixed effect model results for global brain age estimates (fullbrain) and for each region, where variables are displayed with corresponding fixed effect estimates ( $\beta$ ), (standard error), t-statistic, and FDR corrected $P$ value. “Group” levels= WLC (reference, N=139) and AAS (N=166). \* p-value (uncorrected) <.05, \*\* p-value (uncorrected) <.01, \*\*\* p-value (uncorrected) <.001.

When including an interaction term between group (and subgroups of AAS) and time, few significant main effects were found (table S4-S8). One global BAG model survived FDR correction and revealed significant group by time interaction, indicating accelerate aging in users with ≥10 years of use compared to WLC (β (305) = 3.68, t=3.06, p_FDR_=.024, not displayed in Table S8). The longitudinal findings are depicted in Figure 2 for global brain age gap.

**Figure 2.**
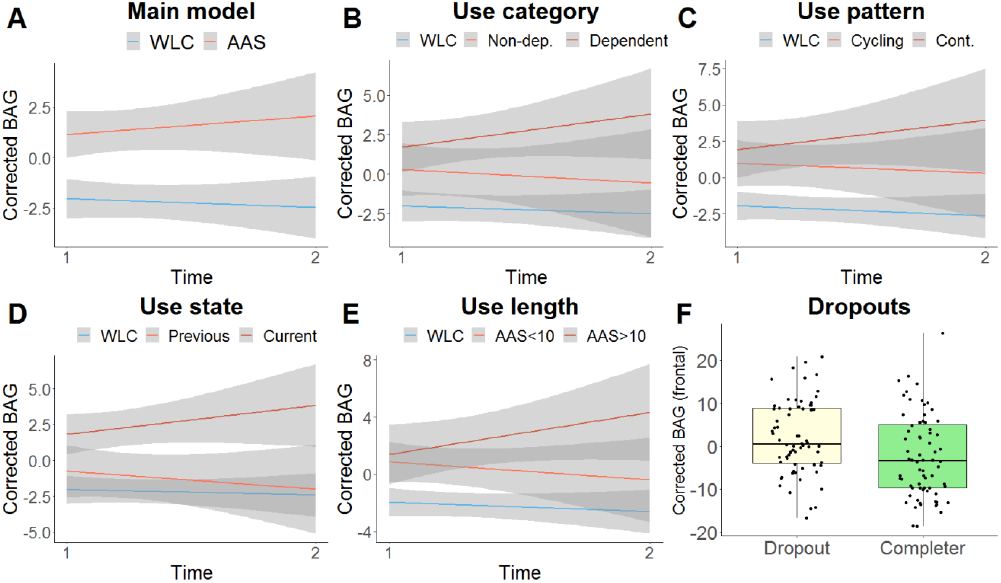
Brain age gap in subgroups. Panel A-E shows group*time (x-axis) interaction for corrected brain age gap (BAG) (y-axis) of subgroups of participants with two scans approximately 3.5 years apart. Fitted lines made with lme-derived predicted values. Shaded gray areas represent CI of 95%. Panel F shows box-plot of corrected BAG at baseline for participants who completed or dropped out of the study. Horizontal lines represents median of sample. Abbreviations: BAG; brain age gap, WLC; weightlifting controls, AAS; anabolic androgenic steroids, Non-dep; non-dependent, Cont; continuous use.

### Sensitivity analyses

Sensitivity analyses revealed that the main effect of group remained significant for the global BAG when IQ, non-AAS substance use and depression were included as fixed effects in the model (Table 3). Frontal and subcortical BAG differences were found at an FDR corrected threshold of p<.05, whereas group differences for the temporal, insula, cingulate and occipital model were no longer significant when adding covariates. Further, the sensitivity analyzes applying stricter inclusion criteria omitting AAS users with <2 years of AAS-use revealed significant main effects of group, with higher BAG for AAS users in all but the subcortical models (Table S9).

**Table 3:**
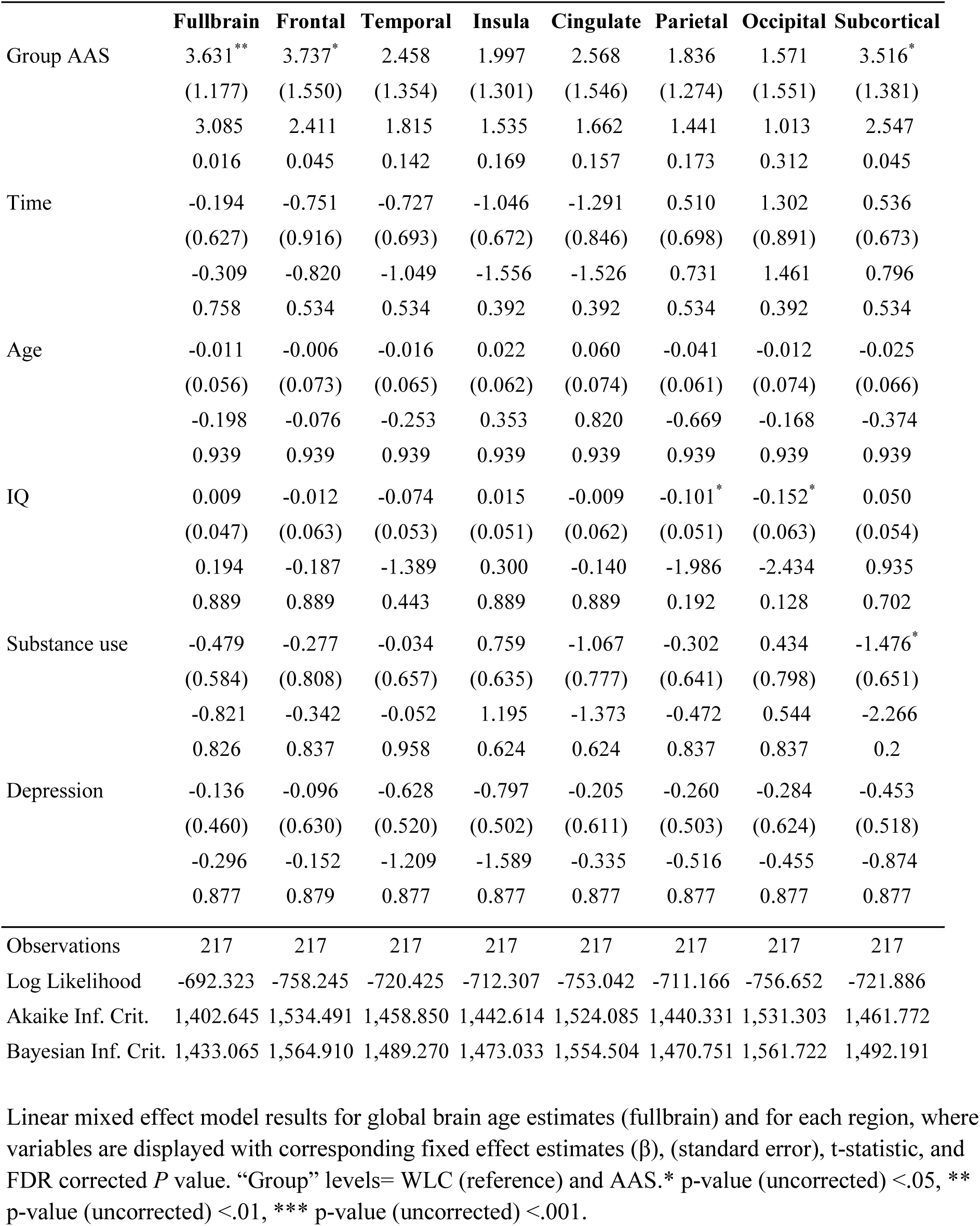
Main model with covariates.

Sensitivity analyses with WLC and subgroups of AAS users revealed significant main effects of use category and higher BAG in dependent compared to WLC for all regions, whereas non-dependent AAS users showed no significant differences from WLC (Table S10), significant main effects of use pattern and higher global BAG and for some regional models in both cyclic and continuous AAS administration compared to WLC (Table S11), and significant main effects of use state with ongoing use, where current AAS users had significantly higher BAG in most regions compared to WLC (Table S12). Previous users (>6 months since last use) were not significantly different from WLC, although differences were seen at an uncorrected significance level for some models including the fullbrain measure ((302) = 2.55, t=2.24, p=.03). Lastly, splitting the groups into shorter (<10 years) versus longer (≥10 years) history of AAS use revealed significant main effects of use length and higher BAG compared to WLC for the fullbrain model and some subregions, with most pronounced differences seen with longer exposure (Table S13).

### BAG associated with dropout

56.7% of the WLC and 46.3% of the AAS users from TPI participated at TPII. Frontal and cingulate BAG at baseline was significantly higher in participants who dropped out of the study compared to those with complete longitudinal data, whereas no significant differences were seen for other regions and the global BAG (Table 4).

**Table 4.**
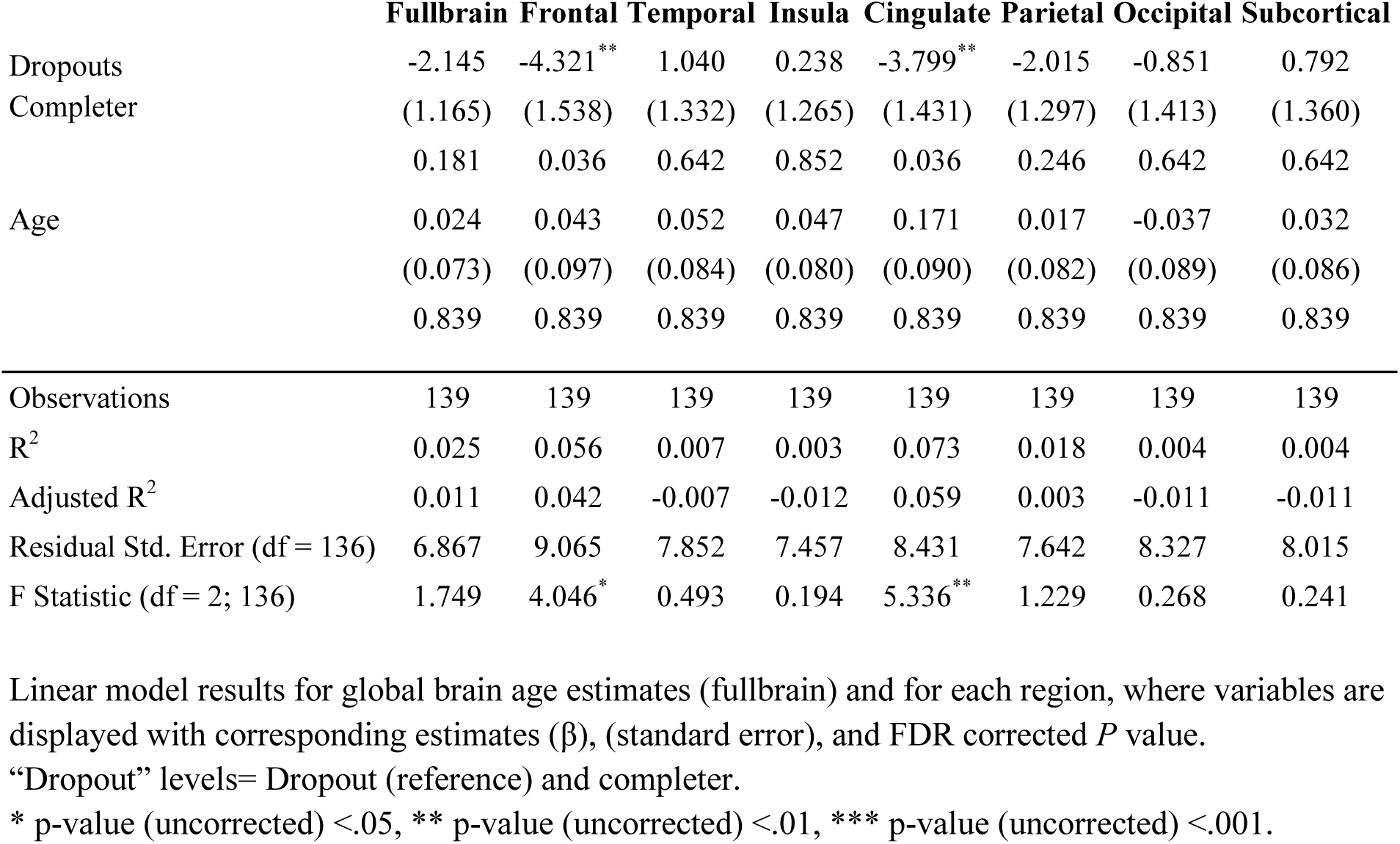
Baseline brain age gap for dropouts (after TP1) and completers across groups.

## Discussion

Accumulating evidence suggests that prolonged AAS use has adverse effects on the brain (12, 36, 37, 39, 42, 43). Using brain scans and brain age prediction based on an independent training set we found evidence of higher relative global, frontal, temporal, occipital and insular brain age in 130 male AAS users compared to 99 male WLC. Further, among AAS users we found that long-term use and dependence were associated with higher relative brain age. Longitudinal analysis revealed no evidence of accelerated BAG over time in the overall AAS group, however AAS users with more than 10 years of AAS exposure showed accelerated aging compared to WLC, with a significant increase in BAG between the time points in this subgroup. These findings suggest that long-term high dose AAS use may have adverse effect on brain aging, potentially linked to dependency and exaggerated use of AAS.

### AAS use associated with apparent brain aging

More evident brain aging in long term AAS users is consistent with *in vitro* studies suggesting that various sorts of AAS might have neurotoxic effects (19-24), and recent findings of impaired cognitive performance (12, 39, 40), smaller brain volumes (37), and metabolites abnormalities (36), associated with long-term AAS use. Older appearing brains in AAS-dependent compared to non-dependent users is consistent with a mega-analyses pooling data from 23 cohorts, suggesting that dependency shares a common neural substrate across a range of substances, indicating smaller brain volumes and thinner cortex in dependent relative to non-dependent individuals (75). The group difference in global BAG suggests widespread effects, although regional models revealed significant differences in several regions, most pronounced frontally. Interestingly, the insula and part of the frontal cortex have been implicated in substance dependence (76-79), and our findings align with structural MRI studies showing reduced insula and frontal gray matter volume in drug users (75, 80).

AAS dependence, current use and longer history of AAS of use were associated with higher BAG. The apparent difference in BAG between past and current AAS users should be regarded with caution, and could be confounded by past users having used AAS for considerably shorter time than current users. The links between AAS use and brain aging are likely complex and reflecting individual vulnerability, properties with the compounds being administered and potential links to medically induced side-effects. In line with this, users with ≥10 years of AAS exposure or AAS dependency, which is characterized by more exaggerated use, the presence of psychological and/or medical side-effects, and continued use despite negative impact on life (1, 15, 40), showed the most prominent accelerated aging over time compared to WLC.

Moreover, we found that AAS users who had dropped out of the study after TPI had older appearing brains in frontal and cingulate regions, compared to those who completed. Hence with a dropout rate of 49% in the total sample and 54% in the AAS user group, it is likely that our longitudinal findings are biased.

Some limitations should be noted. Whereas we included both cross-sectional and longitudinal data, the high drop-out rate and non-random attrition might have limited the generalizability of the longitudinal models. This is in line with previous longitudinal studies of brain aging and dementia, showing that study drop-out is associated with past worse executive and memory functioning (81) and MRI findings suggestive of higher future dementia risk (82). Furthermore, the age distribution of the sample is not optimal as middle and old age is not well represented, and the findings are likely better generalizable to the young adults rather than older male AAS populations. Moreover, while the total sample size is relatively large considering barriers of recruiting participants when studying clandestine and illegal behaviors, our sensitivity analyses resulted in small subpopulations and estimates with high uncertainty. For instance, while past users did not differ from WLC, which could suggest part or full recovery after ceasing AAS use, larger follow-up studies of past users covering a wide age-range are warranted to make plausible conclusions about recovery. It will also be important to study a potential link between long-term AAS use on white matter measures, e.g. measured using diffusion MRI, and, given the strong vascular effects of AAS (27, 83, 84), with slowly progressive vascular pathology such as small vessel disease.

The group differences could not be explained by general cognitive abilities, depression or non-AAS substance use. Still, AAS use is commonly combined with a variety of drugs, such as aromatase inhibitors, human chorionic gonadotropin (hCG), tamoxifen, 5-α-reductase inhibitors, growth hormone (GH), insulin-like growth factor (IGF-1), dietary supplements, as well as narcotics and stimulants (85). In addition, the intricate administration pattern of AAS typically includes different doses and stacking of multiple classes of AAS with different molecular and cellular effects (86). Such complexity makes it extremely difficult to distinguish the unique contributions of single factors on measures of brain health and behavior. Moreover, a range of psychological and medical effects linked to AAS use might influence brain health (15). Hence, future interdisciplinary studies are needed in order to better understand mechanisms linking AAS use and brain aging.

Conclusively, in line with mounting evidence of adverse health effects of AAS use, using brain age prediction we found evidence of increased apparent brain aging in long-term high-dose AAS users, seemingly linked to dependency and exaggerated use of AAS.

## Data Availability

Data sets for model training were partly gathered from public Resources (Links to websites are listed in Table S1). We do not have the ethical permission to share data on our test sample (AAS users and weightlifting controls).

## Acknowledgments

This research was funded by grants 2013087, 2016049, 2017025 and 2018075 (Dr Bjørnebekk) from the South-Eastern Norway Regional Health Authority, and internal research grants from the Division on Mental Health and Addiction (Dr Bjørnebekk). L.T.W. is funded by the European Research Council under the European Union’s Horizon 2020 research and innovation program (ERC Starting Grant 802998), the Research Council of Norway (249795, 273345, 298646), the South-East Norway Regional Health Authority (2019101), and the Department of Psychology, University of Oslo. T.K. is funded by the Research Council of Norway (276082).

Data sets for model training were partly gathered from public resources. <u>CAMCAM:</u> Data collection and sharing for this project was provided by the Cambridge Centre for Ageing and Neuroscience (CamCAN; https://camcan-archive.mrc-cbu.cam.ac.uk/dataaccess/). CamCAN funding was provided by the UK Biotechnology and Biological Sciences Research Council (grant number BB/H008217/1), together with support from the UK Medical Research Council and University of Cambridge, UK. <u>DLBC:</u> Data sets were obtained from http://fcon_1000.projects.nitrc.org/ <u>HPC:</u> Data were provided [in part] by the Human Connectome Project, WU-Minn Consortium (Principal Investigators: David Van Essen and Kamil Ugurbil; 1U54MH091657) funded by the 16 NIH Institutes and Centers that support the NIH Blueprint for Neuroscience Research; and by the McDonnell Center for Systems Neuroscience at Washington University”. For details, see https://www.humanconnectome.org/study/hcp-young-adult/document/hcp-citations. <u>DS000222:</u> Data sets were obtained from the OpenfMRI database https://openfmri.org/ <u>IXI</u>: Data sets were obtained from http://brain-development.org/ixi-dataset/ <u>OASIS</u>: Data sets were obtained from http://www.oasis-brains.org/. The study was supported by grants P50 AG05681, P01 AG03991, R01 AG021910, P50 MH071616, U24 RR021382, R01 MH56584.40 <u>SALD</u>: Data sets were obtained from http://fcon_1000.projects.nitrc.org/. <u>STROKEMRI</u>: Data collection in STROKEMRI was supported by the Research Council of Norway (249795, 248238), the South-Eastern Norway Regional Health Authority (2014097, 2015044, 2015073, 2016083), and the Norwegian Extra Foundation for Health and Rehabilitation (2015/FO5146).

*This article was published as a preprint on medRxiv: doi:* https://www.medrxiv.org/content/10.1101/2020.08.19.20177832v1.article-info

## Disclosures

The authors declare that they have no conflict of interest.

The founding organizations had no role in the design or conduct of the study; in the collection, analysis, or interpretation of data; or in the preparation, review, or approval of the manuscript.

---

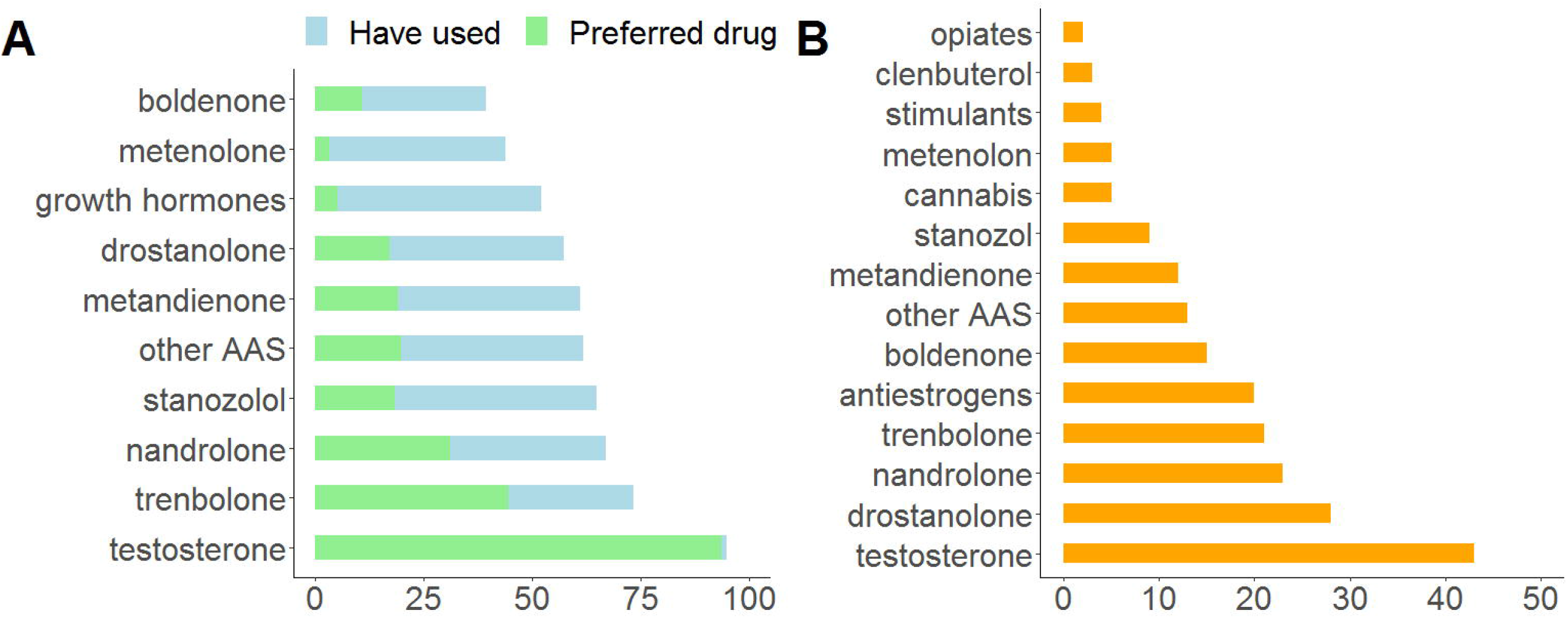

## Notes

### Competing Interest Statement

The authors have declared no competing interest.

### Clinical Trial

The study is not a clinical trial or intervention study

### Funding Statement

This research was funded by grants 2013087, 2016049, 2017025 and 2018075 (Dr Bjornebekk) from the South-Eastern Norway Regional Health Authority, and internal research grants from the Division on Mental Health and Addiction (Dr Bjornebekk). L.T.W. is funded by the European Research Council under the European Unions Horizon 2020 research and innovation program (ERC Starting Grant 802998), the Research Council of Norway (249795, 273345, 298646), the South-East Norway Regional Health Authority (2019101), and the Department of Psychology, University of Oslo. T.K. is funded by the Research Council of Norway (276082).

### Author Declarations

The study was approved by the Regional Committees for Medical and Health Research Ethics South East Norway (REC) (2013/601)

## References

1. Kanayama G, Hudson JI, Pope HG, Jr. (2009): Features of men with anabolic-androgenic steroid dependence: A comparison with nondependent AAS users and with AAS nonusers. Drug and alcohol dependence. 102:130–137.

2. Bhasin S, Storer TW, Berman N, Callegari C, Clevenger B, Phillips J, et al. (1996): The effects of supraphysiologic doses of testosterone on muscle size and strength in normal men. The New England journal of medicine. 335:1–7.

3. Pope HG, Jr., Kouri EM, Hudson JI (2000): Effects of supraphysiologic doses of testosterone on mood and aggression in normal men: a randomized controlled trial. Archives of general psychiatry. 57:133-140; discussion 155-136.

4. Thiblin I, Runeson B, Rajs J (1999): Anabolic androgenic steroids and suicide. Annals of clinical psychiatry : official journal of the American Academy of Clinical Psychiatrists. 11:223–231.

5. Basaria S, Wahlstrom JT, Dobs AS (2001): Clinical review 138: Anabolic-androgenic steroid therapy in the treatment of chronic diseases. The Journal of clinical endocrinology and metabolism. 86:5108–5117.

6. Shahidi NT (2001): A review of the chemistry, biological action, and clinical applications of anabolic-androgenic steroids. Clin Ther. 23:1355–1390.

7. Pomerantz SM, Fox TO, Sholl SA, Vito CC, Goy RW (1985): Androgen and estrogen receptors in fetal rhesus monkey brain and anterior pituitary. Endocrinology. 116:83–89.

8. Simerly RB, Chang C, Muramatsu M, Swanson LW (1990): Distribution of androgen and estrogen receptor mRNA-containing cells in the rat brain: an in situ hybridization study. The Journal of comparative neurology. 294:76–95.

9. Mitra SW, Hoskin E, Yudkovitz J, Pear L, Wilkinson HA, Hayashi S, et al. (2003): Immunolocalization of estrogen receptor beta in the mouse brain: comparison with estrogen receptor alpha. Endocrinology. 144:2055–2067.

10. Reyes-Fuentes A, Veldhuis JD (1993): Neuroendocrine physiology of the normal male gonadal axis. Endocrinol Metab Clin North Am. 22:93–124.

11. Pope HG, Jr., Wood RI, Rogol A, Nyberg F, Bowers L, Bhasin S (2014): Adverse health consequences of performance-enhancing drugs: an Endocrine Society scientific statement. Endocr Rev. 35:341–375.

12. Bjørnebekk A, Westlye LT, Walhovd KB, Jørstad ML, Sundseth ØØ, Fjell AM (2019): Cognitive performance and structural brain correlates in long-term anabolic-androgenic steroid exposed and nonexposed weightlifters. Neuropsychology. 33:547–559.

13. Brower KJ (2002): Anabolic steroid abuse and dependence. Current psychiatry reports. 4:377–387.

14. Ip EJ, Barnett MJ, Tenerowicz MJ, Perry PJ (2011): The Anabolic 500 survey: characteristics of male users versus nonusers of anabolic-androgenic steroids for strength training. Pharmacotherapy. 31:757–766.

15. Kanayama G, Brower KJ, Wood RI, Hudson JI, Pope HG, Jr. (2009): Anabolic-androgenic steroid dependence: an emerging disorder. Addiction. 104:1966–1978.

16. Chandler M, McVeigh J (2013): Steroids and Image Enhancing Drugs 2013 Survey Results. Centre for Public Health: Liverpool John Moores University.

17. Brower KJ (1997): Withdrawal from anabolic steroids. Current therapy in endocrinology and metabolism. 6:338–343.

18. Kashkin KB, Kleber HD (1989): Hooked on hormones? An anabolic steroid addiction hypothesis. JAMA : the journal of the American Medical Association. 262:3166–3170.

19. Basile JR, Binmadi NO, Zhou H, Yang YH, Paoli A, Proia P (2013): Supraphysiological doses of performance enhancing anabolic-androgenic steroids exert direct toxic effects on neuron-like cells. Front Cell Neurosci. 7:69.

20. Caraci F, Pistara V, Corsaro A, Tomasello F, Giuffrida ML, Sortino MA, et al. (2011): Neurotoxic properties of the anabolic androgenic steroids nandrolone and methandrostenolone in primary neuronal cultures. Journal of neuroscience research. 89:592–600.

21. Estrada M, Varshney A, Ehrlich BE (2006): Elevated testosterone induces apoptosis in neuronal cells. J Biol Chem. 281:25492–25501.

22. Westlye ET, Lundervold A, Rootwelt H, Lundervold AJ, Westlye LT (2011): Increased hippocampal default mode synchronization during rest in middle-aged and elderly APOE epsilon4 carriers: relationships with memory performance. The Journal of neuroscience : the official journal of the Society for Neuroscience. 31:7775–7783.

23. Zelleroth S, Nylander E, Nyberg F, Gronbladh A, Hallberg M (2019): Toxic Impact of Anabolic Androgenic Steroids in Primary Rat Cortical Cell Cultures. Neuroscience. 397:172–183.

24. Orlando R, Caruso A, Molinaro G, Motolese M, Matrisciano F, Togna G, et al. (2007): Nanomolar concentrations of anabolic-androgenic steroids amplify excitotoxic neuronal death in mixed mouse cortical cultures. Brain research. 1165:21–29.

25. Hammond J, Le Q, Goodyer C, Gelfand M, Trifiro M, LeBlanc A (2001): Testosterone-mediated neuroprotection through the androgen receptor in human primary neurons. J Neurochem. 77:1319–1326.

26. Nguyen TV, Yao M, Pike CJ (2005): Androgens activate mitogen-activated protein kinase signaling: role in neuroprotection. J Neurochem. 94:1639–1651.

27. Baggish AL, Weiner RB, Kanayama G, Hudson JI, Lu MT, Hoffmann U, et al. (2017): Cardiovascular Toxicity of Illicit Anabolic-Androgenic Steroid Use. Circulation. 135:1991–2002.

28. Nottin S, Nguyen LD, Terbah M, Obert P (2006): Cardiovascular effects of androgenic anabolic steroids in male bodybuilders determined by tissue Doppler imaging. The American journal of cardiology. 97:912–915.

29. Kanayama G, Hudson JI, DeLuca J, Isaacs S, Baggish A, Weiner R, et al. (2015): Prolonged hypogonadism in males following withdrawal from anabolic-androgenic steroids: an under-recognized problem. Addiction. 110:823–831.

30. Rasmussen JJ, Selmer C, Ostergren PB, Pedersen KB, Schou M, Gustafsson F, et al. (2016): Former Abusers of Anabolic Androgenic Steroids Exhibit Decreased Testosterone Levels and Hypogonadal Symptoms Years after Cessation: A Case-Control Study. PloS one. 11:e0161208.

31. Gheshlaghi F, Piri-Ardakani MR, Masoumi GR, Behjati M, Paydar P (2015): Cardiovascular manifestations of anabolic steroids in association with demographic variables in body building athletes. J Res Med Sci. 20:165–168.

32. Rasmussen JJ, Schou M, Selmer C, Johansen ML, Gustafsson F, Frystyk J, et al. (2017): Insulin sensitivity in relation to fat distribution and plasma adipocytokines among abusers of anabolic androgenic steroids. Clin Endocrinol (Oxf).

33. Modlinski R, Fields KB (2006): The effect of anabolic steroids on the gastrointestinal system, kidneys, and adrenal glands. Curr Sports Med Rep. 5:104–109.

34. de Lange AG, Anaturk M, Suri S, Kaufmann T, Cole JH, Griffanti L, et al. (2020): Multimodal brain-age prediction and cardiovascular risk: The Whitehall II MRI sub-study. NeuroImage. 222:117292.

35. Gorelick PB, Scuteri A, Black SE, Decarli C, Greenberg SM, Iadecola C, et al. (2011): Vascular contributions to cognitive impairment and dementia: a statement for healthcare professionals from the american heart association/american stroke association. Stroke. 42:2672–2713.

36. Kaufman MJ, Janes AC, Hudson JI, Brennan BP, Kanayama G, Kerrigan AR, et al. (2015): Brain and cognition abnormalities in long-term anabolic-androgenic steroid users. Drug and alcohol dependence. 1:47–56.

37. Bjørnebekk A, Walhovd KB, Jørstad ML, Due-Tønnessen P, Hullstein IR, Fjell AM (2017): Structural Brain Imaging of Long-Term Anabolic-Androgenic Steroid Users and Nonusing Weightlifters. Biological psychiatry. 82:294–302.

38. Westlye LT, Kaufmann T, Alnaes D, Hullstein IR, Bjørnebekk A (2017): Brain connectivity aberrations in anabolic-androgenic steroid users. Neuroimage Clin. 13:62–69.

39. Kanayama G, Kean J, Hudson JI, Pope HG, Jr. (2013): Cognitive deficits in long-term anabolic-androgenic steroid users. Drug and alcohol dependence. 130:208–214.

40. Hauger LE, Westlye LT, Bjørnebekk A (2020): Anabolic androgenic steroid dependence is associated with executive dysfunction. Drug and alcohol dependence. 208:107874.

41. Heffernan TM, Battersby L, Bishop P, O’Neill TS (2015): Everyday Memory Deficits Associated with Anabolic-Androgenic Steroid Use in Regular Gymnasium Users. The open psychiatry journal. 9:1–6.

42. Kaufman MJ, Kanayama G, Hudson JI, Pope HG, Jr. (2019): Supraphysiologic-dose anabolic-androgenic steroid use: A risk factor for dementia? Neurosci Biobehav Rev. 100:180–207.

43. Pomara C, Neri M, Bello S, Fiore C, Riezzo I, Turillazzi E (2015): Neurotoxicity by Synthetic Androgen Steroids: Oxidative Stress, Apoptosis, and Neuropathology: A Review. Curr Neuropharmacol. 13:132–145.

44. Hall RC, Chapman MJ (2005): Psychiatric complications of anabolic steroid abuse. Psychosomatics. 46:285–290.

45. Hauger LE, Sagoe D, Vaskinn A, Arnevik EA, Leknes S, Jørstad ML, et al. (2019): Anabolic androgenic steroid dependence is associated with impaired emotion recognition. Psychopharmacology (Berl). 236:2667–2676.

46. Etkin A (2019): A Reckoning and Research Agenda for Neuroimaging in Psychiatry. The American journal of psychiatry. 176:507–511.

47. Dosenbach NU, Nardos B, Cohen AL, Fair DA, Power JD, Church JA, et al. (2010): Prediction of individual brain maturity using fMRI. Science. 329:1358–1361.

48. Franke K, Ziegler G, Kloppel S, Gaser C, Alzheimer’s Disease Neuroimaging I (2010): Estimating the age of healthy subjects from T1-weighted MRI scans using kernel methods: exploring the influence of various parameters. NeuroImage. 50:883–892.

49. Cole JH, Franke K (2017): Predicting Age Using Neuroimaging: Innovative Brain Ageing Biomarkers. Trends in neurosciences. 40:681–690.

50. Cole JH, Poudel RPK, Tsagkrasoulis D, Caan MWA, Steves C, Spector TD, et al. (2017): Predicting brain age with deep learning from raw imaging data results in a reliable and heritable biomarker. NeuroImage. 163:115–124.

51. Liem F, Varoquaux G, Kynast J, Beyer F, Kharabian Masouleh S, Huntenburg JM, et al. (2017): Predicting brain-age from multimodal imaging data captures cognitive impairment. NeuroImage. 148:179–188.

52. Cole JH, Ritchie SJ, Bastin ME, Valdes Hernandez MC, Munoz Maniega S, Royle N, et al. (2018): Brain age predicts mortality. Mol Psychiatry. 23:1385–1392.

53. Lebedeva AK, Westman E, Borza T, Beyer MK, Engedal K, Aarsland D, et al. (2017): MRI-Based Classification Models in Prediction of Mild Cognitive Impairment and Dementia in Late-Life Depression. Frontiers in aging neuroscience. 9:13.

54. Kaufmann T, van der Meer D, Doan NT, Schwarz E, Lund MJ, Agartz I, et al. (2019): Common brain disorders are associated with heritable patterns of apparent aging of the brain. Nature neuroscience. 22:1617–1623.

55. Steffener J, Habeck C, O’Shea D, Razlighi Q, Bherer L, Stern Y (2016): Differences between chronological and brain age are related to education and self-reported physical activity. Neurobiology of aging. 40:138–144.

56. Ersche KD, Jones PS, Williams GB, Robbins TW, Bullmore ET (2013): Cocaine dependence: a fast-track for brain ageing? Mol Psychiatry. 18:134–135.

57. Nakama H, Chang L, Fein G, Shimotsu R, Jiang CS, Ernst T (2011): Methamphetamine users show greater than normal age-related cortical gray matter loss. Addiction. 106:1474–1483.

58. Guggenmos M, Schmack K, Sekutowicz M, Garbusow M, Sebold M, Sommer C, et al. (2017): Quantitative neurobiological evidence for accelerated brain aging in alcohol dependence. Translational psychiatry. 7:1279.

59. Bachi K, Sierra S, Volkow ND, Goldstein RZ, Alia-Klein N (2017): Is biological aging accelerated in drug addiction? Curr Opin Behav Sci. 13:34–39.

60. de Lange AG, Barth C, Kaufmann T, Maximov, II, van der Meer D, Agartz I, et al. (2020): Women’s brain aging: Effects of sex-hormone exposure, pregnancies, and genetic risk for Alzheimer’s disease. Human brain mapping. 41:5141–5150.

61. Dale AM, Fischl B, Sereno MI (1999): Cortical surface-based analysis. I. Segmentation and surface reconstruction. NeuroImage. 9:179–194.

62. Glasser MF, Coalson TS, Robinson EC, Hacker CD, Harwell J, Yacoub E, et al. (2016): A multi-modal parcellation of human cerebral cortex. Nature. 536:171–178.

63. Richard G, Kolskar K, Sanders AM, Kaufmann T, Petersen A, Doan NT, et al. (2018): Assessing distinct patterns of cognitive aging using tissue-specific brain age prediction based on diffusion tensor imaging and brain morphometry. PeerJ. 6:e5908.

64. Le TT, Kuplicki RT, McKinney BA, Yeh HW, Thompson WK, Paulus MP, et al. (2018): A Nonlinear Simulation Framework Supports Adjusting for Age When Analyzing BrainAGE. Frontiers in aging neuroscience. 10:317.

65. de Lange AG, Cole JH (2020): Commentary: Correction procedures in brain-age prediction. Neuroimage Clin. 26:102229.

66. Saunders JB, Aasland OG, Babor TF, de la Fuente JR, Grant M (1993): Development of the Alcohol Use Disorders Identification Test (AUDIT): WHO Collaborative Project on Early Detection of Persons with Harmful Alcohol Consumption--II. Addiction. 88:791–804.

67. Berman AH, Bergman H, Palmstierna T, Schlyter F (2005): Evaluation of the Drug Use Disorders Identification Test (DUDIT) in criminal justice and detoxification settings and in a Swedish population sample. Eur Addict Res. 11:22–31.

68. Hauger LE, Westlye LT, Fjell AM, Walhovd KB, Bjornebekk A (2019): Structural brain characteristics of anabolic-androgenic steroid dependence in men. Addiction. 114:1405–1415.

69. Pope HG, Jr., Katz DL (1994): Psychiatric and medical effects of anabolic-androgenic steroid use. A controlled study of 160 athletes. Archives of general psychiatry. 51:375–382.

70. Wechsler D (1999): Wechsler abbreviated scale of intelligens. San Antonio, TX: The Psychological Corporation.

71. Hullstein IR, Malerod-Fjeld H, Dehnes Y, Hemmersbach P (2015): Black market products confiscated in Norway 2011-2014 compared to analytical findings in urine samples. Drug Test Anal. 7:1025–1029.

72. Team RC (2014): R: A Language and Environment for Statistical Computing. R Foundation for Statistical Computing.

73. Bates D, Mächler M, Bolker B, Walker S (2015): Fitting Linear Mixed-Effects Models Using lme4. J Journal of Statistical Software. 67:48.

74. Benjamini Y, Hochberg Y (1995): Controlling The False Discovery Rate - A Practical And Powerful Approach To Multiple Testing. J Royal Statist Soc, Series B. 57:289–300.

75. Mackey S, Allgaier N, Chaarani B, Spechler P, Orr C, Bunn J, et al. (2019): Mega-Analysis of Gray Matter Volume in Substance Dependence: General and Substance-Specific Regional Effects. The American journal of psychiatry. 176:119–128.

76. Everitt BJ, Robbins TW (2005): Neural systems of reinforcement for drug addiction: from actions to habits to compulsion. Nature neuroscience. 8:1481–1489.

77. Goldstein RZ, Craig AD, Bechara A, Garavan H, Childress AR, Paulus MP, et al. (2009): The neurocircuitry of impaired insight in drug addiction. Trends in cognitive sciences. 13:372–380.

78. Koob GF, Volkow ND (2010): Neurocircuitry of addiction. Neuropsychopharmacology : official publication of the American College of Neuropsychopharmacology. 35:217–238.

79. Droutman V, Read SJ, Bechara A (2015): Revisiting the role of the insula in addiction. Trends in cognitive sciences. 19:414–420.

80. Ersche KD, Williams GB, Robbins TW, Bullmore ET (2013): Meta-analysis of structural brain abnormalities associated with stimulant drug dependence and neuroimaging of addiction vulnerability and resilience. Curr Opin Neurobiol. 23:615–624.

81. Sünkel U, Heinzel S, Thaler A-K, Metzger F, Eschweiler G, Liepelt-Scarfone I, et al. (2016): Characteristics of dropouts in a longitudinal study of individuals at risk for a neurodegenerative disorder. MDS Congress. Berlin.

82. Glymour MM, Chene G, Tzourio C, Dufouil C (2012): Brain MRI markers and dropout in a longitudinal study of cognitive aging: the Three-City Dijon Study. Neurology. 79:1340–1348.

83. Rasmussen JJ, Schou M, Madsen PL, Selmer C, Johansen ML, Hovind P, et al. (2018): Increased blood pressure and aortic stiffness among abusers of anabolic androgenic steroids: potential effect of suppressed natriuretic peptides in plasma? J Hypertens. 36:277–285.

84. Vanberg P, Atar D (2010): Androgenic anabolic steroid abuse and the cardiovascular system. Handbook of experimental pharmacology.411–457.

85. Sagoe D, McVeigh J, Bjornebekk A, Essilfie MS, Andreassen CS, Pallesen S (2015): Polypharmacy among anabolic-androgenic steroid users: a descriptive metasynthesis. Subst Abuse Treat Prev Policy. 10:12.

86. Penatti CA, Porter DM, Henderson LP (2009): Chronic exposure to anabolic androgenic steroids alters neuronal function in the mammalian forebrain via androgen receptor- and estrogen receptor-mediated mechanisms. The Journal of neuroscience : the official journal of the Society for Neuroscience. 29:12484–12496.

