## Supplemental Materials for "Long-term anabolic androgenic steroid use is associated with deviant brain aging"

**Supplementary Material**

**Methods**

**Table S1.** Age distribution and links to websites of the cohorts included in the training sample.

| Sample | Source | Min | Max | Mean | SD |
|---|---|---|---|---|---|
| CAMCAN | https://camcan-archive.mrc-cbu.cam.ac.uk/dataaccess/ | 18 | 87 | 54.26 | 18.36 |
| DLBS | http://fcon_1000.projects.nitrc.org/ | 20.57 | 88.14 | 53.72 | 20.24 |
| ds000222 | https://openfmri.org/ | 23 | 73 | 47.73 | 20.01 |
| HCP | https://www.humanconnectome.org/ | 22 | 37 | 27.87 | 3.59 |
| IXI | http://brain-development.org/ixi-dataset/32/ | 20.07 | 86.2 | 46.68 | 16.37 |
| OASIS | http://www.oasis-brains.org/ | 18 | 90 | 48.23 | 24.68 |
| SALD | http://fcon_1000.projects.nitrc.org/ | 20 | 80 | 45.3 | 17.41 |
| STROKEMRI | Local sample (L.T.W.) | 20 | 92 | 63.94 | 14.43 |

**Results**

**Table S2.** Comparisons of AAS subgroups on measures providing information regarding the extent of the steroid use.

| *Subgroup comparisons* | | | | | | *95% CI* | | |
|---|---|---|---|---|---|---|---|---|
| | *Mean* | *SD* | *Mean* | *SD* | *t* | *LL* | *UL* | *p* |
| | **AAS non dependent** | | **AAS dependent** | | | | | |
| *Years of AAS use* | 7.3 | 6.2 | 13.0 | 7.8 | -4.564 | -8.1 | -3.2 | <.001 |
| *Debut Age* | 23.9 | 7.0 | 20.8 | 5.1 | 2.791 | 0.9 | 5.2 | <.01 |
| *Weekly dose* | 712.9 | 506.9 | 1204.1 | 631.9 | -4.609 | -702.1 | -280.2 | <.001 |
| | **Current AAS users** | | **Past AAS users** | | | | | .415 |
| *Years of AAS use* | 11.9 | 8.2 | 8.3 | 5.9 | 2.772 | 1.0 | 6.1 | <.01 |
| *Debut Age* | 22.9 | 6.7 | 19.9 | 3.0 | 3.371 | 1.2 | 4.7 | <.001 |
| *Weekly dose* | 969.2 | 593.5 | 1079.2 | 705.6 | -.896 | -353.2 | 133.2 | .372 |
| | **AAS Cycling** | | **AAS Continuous use** | | | | | |
| *Years of AAS use* | 9.8 | 7.2 | 12.0 | 8.7 | -1.472 | -5.1 | 0.8 | .144 |
| *Debut Age* | 21.5 | 5.2 | 21.8 | 6.8 | -.307 | -2.6 | 1.9 | .760 |
| *Weekly dose* | 1036.9 | 627.1 | 1060.1 | 625.2 | -.191 | -263.5 | 217.1 | .849 |

**Table S3.** *Pearson correlation between estimated brain age and chronological age within the training sample and the test sample, and the calculated mean absolute error (MAE) and root mean squared error (RMSE) for the different models.*

| | Training sample | | | Test sample | | |
|---|---|---|---|---|---|---|
| Models | *r* | rmse | MAE | *r* | rmse | MAE |
| *Fullbrain* | .93 | 7.57 | 5.76 | .79 | 7.21 | 5.57 |
| *Occipital* | .76 | 12.94 | 10.05 | .71 | 9.06 | 7.30 |
| *Temporal* | .85 | 10.49 | 8.08 | .74 | 7.96 | 6.16 |
| *Frontal* | .88 | 9.55 | 7.19 | .71 | 9.03 | 7.27 |
| *Parietal* | .85 | 10.43 | 7.92 | .76 | 7.86 | 6.10 |
| *Cingulate* | .78 | 12.61 | 9.79 | .73 | 9.13 | 7.15 |
| *Insula* | .79 | 12.22 | 9.49 | .77 | 7.47 | 5.83 |
| *Subcortical* | .90 | 8.54 | 6.46 | .74 | 8.26 | 6.32 |

**Figure 1. Compounds used among AAS users.**
A) Bar plot of self-reported use of AAS and growth hormones. Compounds are categorized as “have used” if participants report they had ever used the compound, and “preferred drug” if participant reported use of the compound in the last cycle, or rated the compound as top three chosen compounds. B) Analytic findings in urine, including findings of antiestrogens and other substances of abuse, such as opiates.

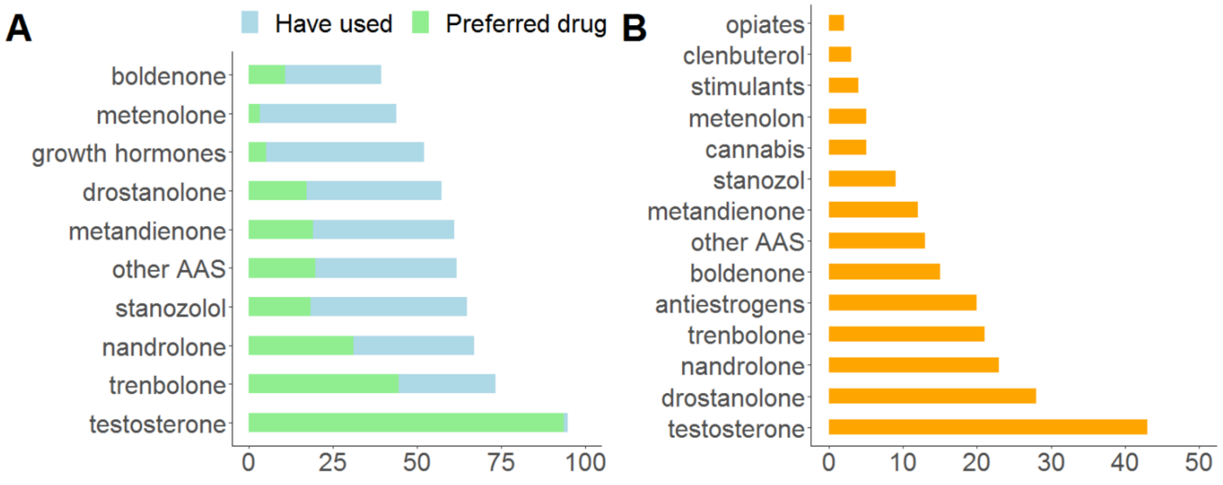

**Table S4: Main model with interaction term (group*time)**

Linear mixed effect model results for global brain age estimates (fullbrain) and for each region, where variables are displayed with corresponding fixed effect estimates (β), (standard error), t-statistic, and p-value(uncorrected). "Group" levels= WLC (reference, N=139), AAS (N=166).
* p-value (uncorrected) <.05, ** p-value (uncorrected) <.01, *** p-value (uncorrected) <.001

| | **Fullbrain** | **Frontal** | **Temporal** | **Insula** | **Cingulate** | **Parietal** | **Occipital** | **Subcortical** |
|---|---|---|---|---|---|---|---|---|
| Group AAS | 1.130 | 2.379 | 2.700 | 1.465 | 2.019 | 0.002 | 2.953 | 0.213 |
| | (1.458) | (2.012) | (1.635) | (1.581) | (1.948) | (1.598) | (1.969) | (1.557) |
| | t = 0.775 | t = 1.182 | t = 1.651 | t = 0.927 | t = 1.036 | t = 0.002 | t = 1.499 | t = 0.137 |
| | p = 0.439 | p = 0.238 | p = 0.099 | p = 0.355 | p = 0.300 | p = 0.999 | p = 0.134 | p = 0.892 |
| Time | -0.612 | -0.326 | -0.795 | -1.291 | -0.676 | -0.813 | 0.901 | 0.109 |
| | (0.683) | (0.974) | (0.759) | (0.747) | (0.927) | (0.742) | (0.953) | (0.694) |
| | t = -0.897 | t = -0.334 | t = -1.048 | t = -1.730 | t = -0.729 | t = -1.096 | t = 0.945 | t = 0.157 |
| | p = 0.370 | p = 0.739 | p = 0.295 | p = 0.084 | p = 0.467 | p = 0.274 | p = 0.345 | p = 0.876 |
| Age | -0.020 | -0.006 | -0.033 | -0.020 | 0.007 | -0.015 | -0.025 | -0.055 |
| | (0.049) | (0.064) | (0.056) | (0.053) | (0.064) | (0.055) | (0.062) | (0.057) |
| | t = -0.405 | t = -0.099 | t = -0.591 | t = -0.379 | t = 0.116 | t = -0.270 | t = -0.395 | t = -0.969 |
| | p = 0.686 | p = 0.922 | p = 0.555 | p = 0.705 | p = 0.908 | p = 0.787 | p = 0.694 | p = 0.333 |
| Group AAS: Time | 1.827 | 1.153 | -0.107 | 0.862 | 0.503 | 1.720 | -0.057 | 1.437 |
| | (0.961) | (1.378) | (1.065) | (1.052) | (1.308) | (1.041) | (1.348) | (0.968) |
| | t = 1.902 | t = 0.837 | t = -0.101 | t = 0.819 | t = 0.384 | t = 1.652 | t = -0.042 | t = 1.485 |
| | p = 0.058 | p = 0.403 | p = 0.920 | p = 0.413 | p = 0.701 | p = 0.099 | p = 0.967 | p = 0.138 |
| Observations | 305 | 305 | 305 | 305 | 305 | 305 | 305 | 305 |
| Log Likelihood | -984.632 | -1,070.367 | -1,022.644 | -1,006.687 | -1,067.315 | -1,015.632 | -1,064.042 | -1,018.158 |
| Akaike Inf. Crit. | 1,983.264 | 2,154.734 | 2,059.288 | 2,027.373 | 2,148.630 | 2,045.264 | 2,142.083 | 2,050.316 |
| Bayesian Inf. Crit. | 2,009.306 | 2,180.776 | 2,085.331 | 2,053.415 | 2,174.672 | 2,071.306 | 2,168.126 | 2,076.359 |

**Table S5: Main model with interaction term (use category*time)**

Linear mixed effect model results for global brain age estimates (fullbrain) and for each region, where variables are displayed with corresponding fixed effect estimates (β), (standard error), t-statistic, and p-value(uncorrected). “Use category” levels= WLC (reference, N=139) non-dependent (N=70), dependent (N=95). * p-value (uncorrected) <.05, ** p-value (uncorrected) <.01, *** p-value (uncorrected) <.001

| | **Fullbrain** | **Frontal** | **Temporal** | **Insula** | **Cingulate** | **Parietal** | **Occipital** | **Subcortical** |
|---|---|---|---|---|---|---|---|---|
| Use category non-dependent | 2.377 | 1.716 | 2.752 | 2.440 | 1.462 | 0.083 | 3.011 | 1.481 |
| | (1.853) | (2.593) | (2.101) | (2.034) | (2.526) | (2.053) | (2.534) | (1.981) |
| | t = 1.283 | t = 0.662 | t = 1.310 | t = 1.200 | t = 0.579 | t = 0.040 | t = 1.188 | t = 0.747 |
| | p = 0.200 | p = 0.509 | p = 0.191 | p = 0.231 | p = 0.563 | p = 0.968 | p = 0.235 | p = 0.455 |
| Use category Dependent | 0.554 | 3.412 | 3.106 | 1.019 | 2.688 | 0.224 | 3.394 | -0.450 |
| | (1.713) | (2.379) | (1.941) | (1.875) | (2.323) | (1.898) | (2.320) | (1.847) |
| | t = 0.324 | t = 1.434 | t = 1.600 | t = 0.543 | t = 1.157 | t = 0.118 | t = 1.463 | t = -0.244 |
| | p = 0.747 | p = 0.152 | p = 0.110 | p = 0.587 | p = 0.248 | p = 0.907 | p = 0.144 | p = 0.808 |
| Time | -0.596 | -0.306 | -0.790 | -1.284 | -0.675 | -0.810 | 0.885 | 0.118 |
| | (0.666) | (0.961) | (0.758) | (0.740) | (0.928) | (0.738) | (0.947) | (0.689) |
| | t = -0.894 | t = -0.318 | t = -1.041 | t = -1.734 | t = -0.728 | t = -1.097 | t = 0.935 | t = 0.172 |
| | p = 0.372 | p = 0.751 | p = 0.298 | p = 0.083 | p = 0.467 | p = 0.273 | p = 0.350 | p = 0.864 |
| Age | -0.024 | -0.011 | -0.037 | -0.022 | 0.006 | -0.017 | -0.028 | -0.058 |
| | (0.049) | (0.063) | (0.055) | (0.052) | (0.063) | (0.055) | (0.060) | (0.057) |
| | t = -0.478 | t = -0.179 | t = -0.667 | t = -0.426 | t = 0.101 | t = -0.306 | t = -0.470 | t = -1.024 |
| | p = 0.633 | p = 0.859 | p = 0.506 | p = 0.671 | p = 0.920 | p = 0.760 | p = 0.639 | p = 0.306 |
| Use category non-dependent: Time | -0.058 | 0.155 | -1.647 | -0.881 | -0.282 | 0.573 | -2.258 | -0.105 |
| | (1.277) | (1.850) | (1.454) | (1.422) | (1.784) | (1.414) | (1.825) | (1.311) |
| | t = -0.045 | t = 0.084 | t = -1.133 | t = -0.619 | t = -0.158 | t = 0.405 | t = -1.237 | t = -0.080 |
| | p = 0.964 | p = 0.934 | p = 0.258 | p = 0.536 | p = 0.875 | p = 0.686 | p = 0.217 | p = 0.937 |
| Use category dependent: Time | 2.846* | 1.354 | 0.530 | 1.793 | 0.688 | 2.194 | 0.900 | 2.327* |
| | (1.123) | (1.621) | (1.278) | (1.248) | (1.564) | (1.243) | (1.598) | (1.157) |
| | t = 2.536 | t = 0.835 | t = 0.415 | t = 1.436 | t = 0.440 | t = 1.765 | t = 0.564 | t = 2.012 |
| | p = 0.012 | p = 0.404 | p = 0.679 | p = 0.151 | p = 0.660 | p = 0.078 | p = 0.574 | p = 0.045 |
| Observations | 304 | 304 | 304 | 304 | 304 | 304 | 304 | 304 |
| Log Likelihood | -977.938 | -1,064.080 | -1,014.442 | -1,000.174 | -1,061.286 | -1,009.395 | -1,051.972 | -1,013.262 |
| Akaike Inf. Crit. | 1,973.877 | 2,146.160 | 2,046.884 | 2,018.348 | 2,140.573 | 2,036.790 | 2,121.943 | 2,044.524 |
| Bayesian Inf. Crit. | 2,007.330 | 2,179.613 | 2,080.337 | 2,051.801 | 2,174.026 | 2,070.244 | 2,155.396 | 2,077.977 |

**Table S6: Main model with interaction term (use pattern*time)**

Linear mixed effect model results for global brain age estimates (fullbrain) and for each region, where variables are displayed with corresponding fixed effect estimates (β), (standard error), t-statistic, and p-value(uncorrected). “Use pattern” levels= WLC (reference, N=139) cycling (N= 92), continuous use (N=57).* p-value (uncorrected) <.05, ** p-value (uncorrected) <.01, *** p-value (uncorrected) <.001

| | **Fullbrain** | **Frontal** | **Temporal** | **Insula** | **Cingulate** | **Parietal** | **Occipital** | **Subcortical** |
|---|---|---|---|---|---|---|---|---|
| Use pattern Cycling | 2.403 | 1.926 | 2.015 | 1.763 | 2.668 | -0.734 | 4.152 | 1.410 |
| | (1.843) | (2.569) | (2.045) | (1.808) | (2.353) | (2.026) | (2.346) | (1.880) |
| | t = 1.304 | t = 0.750 | t = 0.985 | t = 0.976 | t = 1.134 | t = -0.362 | t = 1.769 | t = 0.750 |
| | p = 0.193 | p = 0.454 | p = 0.325 | p = 0.330 | p = 0.257 | p = 0.718 | p = 0.077 | p = 0.454 |
| Use pattern Continuous use | 1.252 | 3.899 | 1.524 | 0.424 | 2.491 | -0.393 | 1.762 | 0.861 |
| | (2.114) | (2.956) | (2.346) | (2.067) | (2.698) | (2.322) | (2.693) | (2.140) |
| | t = 0.592 | t = 1.319 | t = 0.650 | t = 0.205 | t = 0.923 | t = -0.169 | t = 0.654 | t = 0.403 |
| | p = 0.554 | p = 0.188 | p = 0.516 | p = 0.838 | p = 0.356 | p = 0.866 | p = 0.513 | p = 0.688 |
| Time | -0.637 | -0.296 | -0.919 | -1.303* | -0.608 | -0.833 | 0.903 | 0.030 |
| | (0.686) | (0.990) | (0.762) | (0.655) | (0.876) | (0.749) | (0.880) | (0.655) |
| | t = -0.928 | t = -0.299 | t = -1.206 | t = -1.989 | t = -0.694 | t = -1.111 | t = 1.026 | t = 0.046 |
| | p = 0.354 | p = 0.765 | p = 0.228 | p = 0.047 | p = 0.488 | p = 0.267 | p = 0.305 | p = 0.964 |
| Age | -0.013 | -0.016 | 0.001 | -0.015 | -0.010 | -0.010 | -0.006 | -0.035 |
| | (0.051) | (0.066) | (0.057) | (0.053) | (0.065) | (0.057) | (0.064) | (0.059) |
| | t = -0.247 | t = -0.246 | t = 0.026 | t = -0.280 | t = -0.154 | t = -0.180 | t = -0.094 | t = -0.591 |
| | p = 0.805 | p = 0.806 | p = 0.979 | p = 0.780 | p = 0.878 | p = 0.858 | p = 0.926 | p = 0.555 |
| Use pattern Cycling: Time | 0.827 | 1.774 | 0.413 | 0.133 | -0.090 | 2.100 | -0.954 | 0.258 |
| | (1.273) | (1.827) | (1.414) | (1.217) | (1.625) | (1.391) | (1.631) | (1.216) |
| | t = 0.649 | t = 0.971 | t = 0.292 | t = 0.109 | t = -0.055 | t = 1.510 | t = -0.585 | t = 0.212 |
| | p = 0.517 | p = 0.332 | p = 0.771 | p = 0.913 | p = 0.956 | p = 0.132 | p = 0.559 | p = 0.833 |
| Use pattern Continuous use: Time | 1.776 | -0.755 | 1.502 | 2.766* | 0.071 | 2.361 | 1.326 | 1.765 |
| | (1.444) | (2.078) | (1.604) | (1.378) | (1.843) | (1.577) | (1.852) | (1.373) |
| | t = 1.230 | t = -0.363 | t = 0.936 | t = 2.007 | t = 0.038 | t = 1.497 | t = 0.716 | t = 1.285 |
| | p = 0.219 | p = 0.717 | p = 0.350 | p = 0.045 | p = 0.970 | p = 0.135 | p = 0.474 | p = 0.199 |
| Observations | 288 | 288 | 288 | 288 | 288 | 288 | 288 | 288 |
| Log Likelihood | -933.371 | -1,016.096 | -962.860 | -936.957 | -1,003.599 | -962.882 | -1,000.137 | -960.641 |
| Akaike Inf. Crit. | 1,884.742 | 2,050.191 | 1,943.720 | 1,891.915 | 2,025.199 | 1,943.764 | 2,018.274 | 1,939.281 |
| Bayesian Inf. Crit. | 1,917.709 | 2,083.158 | 1,976.686 | 1,924.881 | 2,058.166 | 1,976.731 | 2,051.241 | 1,972.248 |

**Table S7: Main model with interaction term (use state*time)**

Linear mixed effect model results for global brain age estimates (fullbrain) and for each region, where variables are displayed with corresponding fixed effect estimates (β), (standard error), t-statistic, and p-value(uncorrected). “Use state” levels= WLC (reference, N=139) previous user (N=46), current user (N=117).* p-value (uncorrected) <.05, ** p-value (uncorrected) <.01, *** p-value (uncorrected) <.001

| | **Fullbrain** | **Frontal** | **Temporal** | **Insula** | **Cingulate** | **Parietal** | **Occipital** | **Subcortical** |
|---|---|---|---|---|---|---|---|---|
| Use state Previous user | 0.228 | 2.872 | -0.959 | -0.862 | 6.021 | 0.894 | 2.630 | 0.099 |
| | (2.495) | (3.366) | (2.806) | (2.666) | (3.313) | (2.731) | (3.349) | (2.675) |
| | t = 0.091 | t = 0.853 | t = -0.342 | t = -0.323 | t = 1.817 | t = 0.327 | t = 0.785 | t = 0.037 |
| | p = 0.928 | p = 0.394 | p = 0.733 | p = 0.747 | p = 0.070 | p = 0.744 | p = 0.433 | p = 0.971 |
| Use state Current user | 1.103 | 1.555 | 3.711* | 2.438 | 0.706 | 0.006 | 2.891 | -0.126 |
| | (1.643) | (2.232) | (1.846) | (1.759) | (2.191) | (1.793) | (2.232) | (1.747) |
| | t = 0.671 | t = 0.697 | t = 2.010 | t = 1.386 | t = 0.322 | t = 0.003 | t = 1.295 | t = -0.072 |
| | p = 0.503 | p = 0.487 | p = 0.045 | p = 0.166 | p = 0.748 | p = 0.998 | p = 0.196 | p = 0.943 |
| Time | -0.599 | -0.310 | -0.772 | -1.244 | -0.634 | -0.786 | 0.927 | 0.142 |
| | (0.688) | (0.954) | (0.771) | (0.741) | (0.929) | (0.743) | (0.965) | (0.702) |
| | t = -0.871 | t = -0.325 | t = -1.001 | t = -1.678 | t = -0.682 | t = -1.059 | t = 0.960 | t = 0.202 |
| | p = 0.384 | p = 0.746 | p = 0.317 | p = 0.094 | p = 0.495 | p = 0.290 | p = 0.337 | p = 0.840 |
| Age | -0.025 | -0.008 | -0.043 | -0.035 | -0.006 | -0.024 | -0.038 | -0.064 |
| | (0.049) | (0.064) | (0.056) | (0.052) | (0.064) | (0.055) | (0.062) | (0.057) |
| | t = -0.507 | t = -0.121 | t = -0.772 | t = -0.663 | t = -0.091 | t = -0.427 | t = -0.618 | t = -1.128 |
| | p = 0.612 | p = 0.904 | p = 0.441 | p = 0.508 | p = 0.928 | p = 0.670 | p = 0.537 | p = 0.260 |
| Use state Previous user: Time | 1.850 | 0.845 | 1.360 | 1.422 | -2.297 | 1.358 | -0.692 | 0.829 |
| | (1.639) | (2.249) | (1.838) | (1.760) | (2.199) | (1.777) | (2.260) | (1.699) |
| | t = 1.129 | t = 0.376 | t = 0.740 | t = 0.808 | t = -1.044 | t = 0.764 | t = -0.306 | t = 0.488 |
| | p = 0.259 | p = 0.708 | p = 0.460 | p = 0.420 | p = 0.297 | p = 0.445 | p = 0.760 | p = 0.626 |
| Use state Current user: Time | 2.138 | 1.836 | -0.277 | 0.612 | 1.517 | 1.629 | 0.415 | 2.069 |
| | (1.131) | (1.570) | (1.266) | (1.218) | (1.529) | (1.218) | (1.590) | (1.146) |
| | t = 1.891 | t = 1.169 | t = -0.219 | t = 0.503 | t = 0.992 | t = 1.338 | t = 0.261 | t = 1.806 |
| | p = 0.059 | p = 0.243 | p = 0.827 | p = 0.616 | p = 0.322 | p = 0.181 | p = 0.795 | p = 0.071 |
| Observations | 302 | 302 | 302 | 302 | 302 | 302 | 302 | 302 |
| Log Likelihood | -974.856 | -1,058.851 | -1,011.271 | -993.098 | -1,056.472 | -1,005.701 | -1,053.277 | -1,008.197 |
| Akaike Inf. Crit. | 1,967.711 | 2,135.702 | 2,040.543 | 2,004.196 | 2,130.945 | 2,029.402 | 2,124.554 | 2,034.394 |
| Bayesian Inf. Crit. | 2,001.105 | 2,169.096 | 2,073.937 | 2,037.590 | 2,164.339 | 2,062.796 | 2,157.948 | 2,067.788 |

**Table S8: Main model with interaction term (use length*time)**

Linear mixed effect model results for global brain age estimates (fullbrain) and for each region, where variables are displayed with corresponding fixed effect estimates (β), (standard error), t-statistic, and p-value(uncorrected). "Use length" levels= WLC (reference, N=139) AAS<10 (years, N=93), AAS>10 (years, N=73).* p-value (uncorrected) <.05, ** p-value (uncorrected) <.01, *** p-value (uncorrected) <.001

| | **Fullbrain** | **Frontal** | **Temporal** | **Insula** | **Cingulate** | **Parietal** | **Occipital** | **Subcortical** |
|---|---|---|---|---|---|---|---|---|
| Use length AAS< 10 | 3.229 | 4.417 | 4.609* | 2.145 | 2.115 | 0.405 | 4.779* | 1.164 |
| | (1.702) | (2.403) | (1.937) | (1.879) | (2.325) | (1.906) | (2.348) | (1.835) |
| | t = 1.897 | t = 1.838 | t = 2.380 | t = 1.142 | t = 0.910 | t = 0.212 | t = 2.036 | t = 0.634 |
| | p = 0.058 | p = 0.067 | p = 0.018 | p = 0.254 | p = 0.364 | p = 0.832 | p = 0.042 | p = 0.526 |
| Use length AAS> 10 | -0.680 | 0.218 | 0.699 | -0.109 | 2.515 | -0.471 | 1.739 | -0.292 |
| | (1.891) | (2.649) | (2.153) | (2.082) | (2.573) | (2.119) | (2.587) | (2.049) |
| | t = -0.360 | t = 0.082 | t = 0.325 | t = -0.052 | t = 0.978 | t = -0.222 | t = 0.672 | t = -0.142 |
| | p = 0.720 | p = 0.935 | p = 0.746 | p = 0.959 | p = 0.329 | p = 0.824 | p = 0.502 | p = 0.887 |
| Time | -0.579 | -0.328 | -0.798 | -1.362 | -0.627 | -0.818 | 0.953 | 0.149 |
| | (0.665) | (0.969) | (0.755) | (0.742) | (0.924) | (0.743) | (0.948) | (0.694) |
| | t = -0.871 | t = -0.338 | t = -1.057 | t = -1.836 | t = -0.678 | t = -1.100 | t = 1.006 | t = 0.215 |
| | p = 0.384 | p = 0.736 | p = 0.291 | p = 0.067 | p = 0.498 | p = 0.272 | p = 0.315 | p = 0.830 |
| Age | -0.029 | -0.005 | -0.033 | 0.002 | -0.007 | -0.014 | -0.041 | -0.067 |
| | (0.050) | (0.065) | (0.058) | (0.054) | (0.066) | (0.057) | (0.064) | (0.058) |
| | t = -0.570 | t = -0.078 | t = -0.567 | t = 0.029 | t = -0.111 | t = -0.239 | t = -0.650 | t = -1.157 |
| | p = 0.569 | p = 0.938 | p = 0.571 | p = 0.977 | p = 0.912 | p = 0.812 | p = 0.516 | p = 0.248 |
| Use length AAS< 10: Time | -0.280 | -0.592 | -1.760 | 0.949 | -0.045 | 1.411 | -2.184 | 0.227 |
| | (1.181) | (1.731) | (1.341) | (1.321) | (1.647) | (1.321) | (1.693) | (1.225) |
| | t = -0.237 | t = -0.342 | t = -1.313 | t = 0.719 | t = -0.027 | t = 1.068 | t = -1.290 | t = 0.185 |
| | p = 0.813 | p = 0.733 | p = 0.190 | p = 0.473 | p = 0.979 | p = 0.286 | p = 0.197 | p = 0.853 |
| Use length AAS> 10: Time | 3.681** | 2.897 | 1.529 | 1.272 | 0.700 | 2.058 | 1.646 | 2.353 |
| | (1.204) | (1.755) | (1.367) | (1.344) | (1.674) | (1.347) | (1.716) | (1.255) |
| | t = 3.056 | t = 1.651 | t = 1.118 | t = 0.947 | t = 0.418 | t = 1.528 | t = 0.959 | t = 1.874 |
| | p = 0.003 | p = 0.099 | p = 0.264 | p = 0.344 | p = 0.676 | p = 0.127 | p = 0.338 | p = 0.061 |
| Observations | 305 | 305 | 305 | 305 | 305 | 305 | 305 | 305 |
| Log Likelihood | -980.252 | -1,069.013 | -1,020.644 | -1,005.189 | -1,066.711 | -1,015.553 | -1,061.367 | -1,016.389 |
| Akaike Inf. Crit. | 1,978.504 | 2,156.026 | 2,059.288 | 2,028.379 | 2,151.422 | 2,049.107 | 2,140.734 | 2,050.779 |
| Bayesian Inf. Crit. | 2,011.987 | 2,189.509 | 2,092.771 | 2,061.862 | 2,184.905 | 2,082.590 | 2,174.216 | 2,084.261 |

**Table S9: Main model with stricter inclusion bias (>2 years of use for AAS group)**

Linear mixed effect model results for global brain age estimates (fullbrain) and for each region, where variables are displayed with corresponding fixed effect estimates (β), (standard error), t-statistic, and FDR corrected *P* value. "Group" levels= WLC (reference, N=139) and AAS (N=152).
* p-value (uncorrected) <.05, ** p-value (uncorrected) <.01, *** p-value (uncorrected) <.001.

| | **Fullbrain** | **Frontal** | **Temporal** | **Insula** | **Cingulate** | **Parietal** | **Occipital** | **Subcortical** |
|---|---|---|---|---|---|---|---|---|
| Group AAS | 3.526*** | 4.072*** | 2.982** | 2.717** | 3.013* | 2.436* | 3.419** | 2.060 |
| | (0.946) | (1.208) | (1.066) | (1.008) | (1.210) | (1.045) | (1.186) | (1.086) |
| | 3.729 | 3.371 | 2.797 | 2.695 | 2.490 | 2.331 | 2.882 | 1.897 |
| | <.001 | 0.004 | 0.012 | 0.013 | 0.019 | 0.024 | 0.011 | 0.059 |
| Time | 0.304 | 0.486 | -0.878 | -1.032 | -0.395 | -0.110 | 0.853 | 0.812 |
| | (0.533) | (0.733) | (0.589) | (0.567) | (0.716) | (0.575) | (0.732) | (0.544) |
| | 0.570 | 0.663 | -1.491 | -1.818 | -0.551 | -0.191 | 1.165 | 1.492 |
| | 0.666 | 0.666 | 0.371 | 0.371 | 0.666 | 0.849 | 0.494 | 0.371 |
| Age | -0.014 | -0.018 | -0.025 | -0.031 | 0.009 | -0.009 | -0.027 | -0.031 |
| | (0.052) | (0.066) | (0.058) | (0.055) | (0.066) | (0.057) | (0.065) | (0.059) |
| | -0.264 | -0.273 | -0.422 | -0.571 | 0.137 | -0.153 | -0.420 | -0.527 |
| | 0.891 | 0.891 | 0.891 | 0.891 | 0.891 | 0.891 | 0.891 | 0.891 |
| Observations | 291 | 291 | 291 | 291 | 291 | 291 | 291 | 291 |
| Log Likelihood | -944.399 | -1,022.192 | -977.429 | -962.800 | -1,020.396 | -971.297 | -1,018.308 | -974.140 |
| Akaike Inf. Crit. | 1,900.798 | 2,056.385 | 1,966.858 | 1,937.600 | 2,052.793 | 1,954.594 | 2,048.615 | 1,960.280 |
| Bayesian Inf. Crit. | 1,922.838 | 2,078.425 | 1,988.898 | 1,959.640 | 2,074.833 | 1,976.634 | 2,070.655 | 1,982.320 |

**Table S10: Main model with WLC and subsets of AAS group (use category)**
Linear mixed effect model results for global brain age estimates (fullbrain) and for each region, where variables are displayed with corresponding fixed effect estimates (β), (standard error), t-statistic, and FDR corrected *P* value. "Use category" levels= WLC (reference, N=139) non-dependent (N=70), dependent (N=95).* p-value (uncorrected) <.05, ** p-value (uncorrected) <.01, *** p-value (uncorrected) <.001.

| | **Fullbrain** | **Frontal** | **Temporal** | **Insula** | **Cingulate** | **Parietal** | **Occipital** | **Subcortical** |
|---|---|---|---|---|---|---|---|---|
| Use category non-dependent | 2.089 | 1.819 | 0.738 | 1.255 | 1.079 | 0.594 | 0.271 | 1.122 |
| | (1.081) | (1.397) | (1.210) | (1.150) | (1.393) | (1.195) | (1.340) | (1.234) |
| | 1.933 | 1.302 | 0.610 | 1.091 | 0.775 | 0.497 | 0.202 | 0.909 |
| | 0.432 | 0.702 | 0.709 | 0.702 | 0.702 | 0.709 | 0.84 | 0.702 |
| Use category dependent | 4.098*** | 5.082*** | 3.795*** | 3.261** | 3.545** | 2.946** | 4.528*** | 2.478* |
| | (1.006) | (1.295) | (1.127) | (1.069) | (1.293) | (1.113) | (1.239) | (1.157) |
| | 4.073 | 3.925 | 3.367 | 3.050 | 2.742 | 2.646 | 3.654 | 2.141 |
| | <.001 | <.001 | 0.002 | 0.005 | 0.009 | 0.01 | <.001 | 0.033 |
| Time | 0.177 | 0.099 | -0.970 | -0.961 | -0.540 | -0.092 | 0.691 | 0.734 |
| | (0.507) | (0.711) | (0.564) | (0.553) | (0.686) | (0.552) | (0.699) | (0.520) |
| | 0.349 | 0.139 | -1.718 | -1.739 | -0.787 | -0.167 | 0.989 | 1.411 |
| | 0.89 | 0.89 | 0.352 | 0.352 | 0.693 | 0.89 | 0.65 | 0.429 |
| Age | -0.023 | -0.011 | -0.036 | -0.021 | 0.007 | -0.017 | -0.026 | -0.058 |
| | (0.049) | (0.063) | (0.055) | (0.052) | (0.063) | (0.055) | (0.060) | (0.057) |
| | -0.466 | -0.177 | -0.645 | -0.409 | 0.105 | -0.309 | -0.437 | -1.017 |
| | 0.916 | 0.916 | 0.916 | 0.916 | 0.916 | 0.916 | 0.916 | 0.916 |
| Observations | 304 | 304 | 304 | 304 | 304 | 304 | 304 | 304 |
| Log Likelihood | -981.331 | -1,064.436 | -1,015.340 | -1,001.710 | -1,061.422 | -1,010.945 | -1,053.148 | -1,015.442 |
| Akaike Inf. Crit. | 1,976.662 | 2,142.872 | 2,044.680 | 2,017.420 | 2,136.844 | 2,035.890 | 2,120.296 | 2,044.885 |
| Bayesian Inf. Crit. | 2,002.682 | 2,168.891 | 2,070.699 | 2,043.439 | 2,162.863 | 2,061.909 | 2,146.315 | 2,070.904 |

**Table S11: Main model with WLC and subsets of AAS group (use pattern)**

Linear mixed effect model results for global brain age estimates (fullbrain) and for each region, where variables are displayed with corresponding fixed effect estimates (β), (standard error), t-statistic, and FDR corrected *P* value. “Use pattern” levels= WLC (reference, N=139) cycling (N= 92), continuous use (N=57).* p-value (uncorrected) <.05, ** p-value (uncorrected) <.01, *** p-value (uncorrected) <.001

| | Fullbrain | Frontal | Temporal | Insula | Cingulate | Parietal | Occipital | Subcortical |
|---|---|---|---|---|---|---|---|---|
| Use pattern Cycling | 3.360*** | 4.084** | 2.482* | 1.839 | 2.556* | 1.755 | 2.958* | 1.658 |
| | (1.006) | (1.310) | (1.114) | (1.036) | (1.281) | (1.119) | (1.263) | (1.149) |
| | 3.341 | 3.117 | 2.228 | 1.775 | 1.996 | 1.568 | 2.343 | 1.444 |
| | 0.008 | 0.008 | 0.054 | 0.103 | 0.075 | 0.135 | 0.053 | 0.15 |
| Use pattern Continuous use | 3.409** | 2.876 | 3.357** | 3.877*** | 2.586 | 2.410 | 3.476* | 3.064* |
| | (1.138) | (1.496) | (1.260) | (1.166) | (1.449) | (1.267) | (1.431) | (1.277) |
| | 2.995 | 1.923 | 2.664 | 3.325 | 1.784 | 1.903 | 2.428 | 2.399 |
| | 0.012 | 0.066 | 0.021 | 0.008 | 0.076 | 0.066 | 0.027 | 0.027 |
| Time | -0.125 | -0.005 | -0.557 | -0.779 | -0.617 | 0.078 | 0.911 | 0.402 |
| | (0.537) | (0.769) | (0.592) | (0.522) | (0.681) | (0.595) | (0.684) | (0.519) |
| | -0.232 | -0.006 | -0.941 | -1.492 | -0.906 | 0.132 | 1.332 | 0.775 |
| | 0.995 | 0.995 | 0.704 | 0.704 | 0.704 | 0.995 | 0.704 | 0.704 |
| Age | -0.014 | -0.017 | 0.0001 | -0.017 | -0.010 | -0.013 | -0.007 | -0.037 |
| | (0.051) | (0.066) | (0.057) | (0.053) | (0.065) | (0.057) | (0.064) | (0.059) |
| | -0.281 | -0.253 | 0.001 | -0.321 | -0.154 | -0.229 | -0.103 | -0.619 |
| | 0.999 | 0.999 | 0.999 | 0.999 | 0.999 | 0.999 | 0.999 | 0.999 |
| Observations | 288 | 288 | 288 | 288 | 288 | 288 | 288 | 288 |
| Log Likelihood | -934.219 | -1,016.719 | -963.303 | -938.967 | -1,003.602 | -964.766 | -1,000.657 | -961.464 |
| Akaike Inf. Crit. | 1,882.438 | 2,047.437 | 1,940.607 | 1,891.934 | 2,021.204 | 1,943.532 | 2,015.314 | 1,936.929 |
| Bayesian Inf. Crit. | 1,908.078 | 2,073.078 | 1,966.248 | 1,917.575 | 2,046.845 | 1,969.172 | 2,040.954 | 1,962.569 |

**Table S12: Main model with WLC and subsets of AAS group (use state)**

Linear mixed effect model results for global brain age estimates (fullbrain) and for each region, where variables are displayed with corresponding fixed effect estimates (β), (standard error), t-statistic, and FDR corrected *P* value. “Use state” levels= WLC (reference, N=139) previous user (N=46), current user (N=117).* p-value (uncorrected) <.05, ** p-value (uncorrected) <.01, *** p-value (uncorrected) <.001

| | **Fullbrain** | **Frontal** | **Temporal** | **Insula** | **Cingulate** | **Parietal** | **Occipital** | **Subcortical** |
|---|---|---|---|---|---|---|---|---|
| Use state Previous user | 2.553$^{*}$ | 3.862$^{**}$ | 0.921 | 1.025 | 2.783 | 2.614$^{*}$ | 1.676 | 1.037 |
| | (1.140) | (1.493) | (1.280) | (1.204) | (1.486) | (1.261) | (1.460) | (1.281) |
| | 2.240 | 2.587 | 0.720 | 0.851 | 1.872 | 2.072 | 1.148 | 0.810 |
| | 0.104 | 0.08 | 0.472 | 0.472 | 0.124 | 0.104 | 0.403 | 0.472 |
| Use state Current user | 3.553$^{***}$ | 3.702$^{**}$ | 3.298$^{**}$ | 3.084$^{**}$ | 2.645$^{*}$ | 1.865 | 3.417$^{**}$ | 2.295$^{*}$ |
| | (0.959) | (1.244) | (1.081) | (1.012) | (1.244) | (1.071) | (1.209) | (1.102) |
| | 3.704 | 2.977 | 3.050 | 3.047 | 2.126 | 1.742 | 2.826 | 2.082 |
| | <.001 | 0.006 | 0.006 | 0.006 | 0.043 | 0.083 | 0.008 | 0.043 |
| Time | 0.330 | 0.384 | -0.676 | -0.857 | -0.467 | -0.087 | 0.962 | 0.915 |
| | (0.529) | (0.723) | (0.585) | (0.560) | (0.706) | (0.565) | (0.724) | (0.541) |
| | 0.625 | 0.531 | -1.157 | -1.530 | -0.662 | -0.153 | 1.329 | 1.690 |
| | 0.681 | 0.681 | 0.5 | 0.499 | 0.681 | 0.879 | 0.499 | 0.499 |
| Age | -0.025 | -0.007 | -0.044 | -0.035 | -0.003 | -0.024 | -0.037 | -0.064 |
| | (0.050) | (0.064) | (0.056) | (0.052) | (0.064) | (0.055) | (0.062) | (0.057) |
| | -0.506 | -0.112 | -0.795 | -0.678 | -0.041 | -0.425 | -0.602 | -1.109 |
| | 0.895 | 0.967 | 0.895 | 0.895 | 0.967 | 0.895 | 0.895 | 0.895 |
| Observations | 302 | 302 | 302 | 302 | 302 | 302 | 302 | 302 |
| Log Likelihood | -976.973 | -1,059.546 | -1,011.602 | -993.496 | -1,057.705 | -1,006.754 | -1,053.374 | -1,009.845 |
| Akaike Inf. Crit. | 1,967.946 | 2,133.093 | 2,037.203 | 2,000.991 | 2,129.409 | 2,027.508 | 2,120.749 | 2,033.689 |
| Bayesian Inf. Crit. | 1,993.919 | 2,159.066 | 2,063.176 | 2,026.964 | 2,155.382 | 2,053.481 | 2,146.722 | 2,059.662 |

**Table S13: Main model with WLC and subsets of AAS group (use length)**

Linear mixed effect model results for global brain age estimates (fullbrain) and for each region, where variables are displayed with corresponding fixed effect estimates (β), (standard error), t-statistic, and FDR corrected *P* value. “Use length” levels= WLC (reference, N=139) AAS<10 (years, N=93), AAS>10 (years, N=73).* p-value (uncorrected) <.05, ** p-value (uncorrected) <.01, *** p-value (uncorrected) <.001

| | **Fullbrain** | **Frontal** | **Temporal** | **Insula** | **Cingulate** | **Parietal** | **Occipital** | **Subcortical** |
|---|---|---|---|---|---|---|---|---|
| Use length AAS< 10 | 2.660** | 3.576** | 2.424* | 3.191** | 2.021 | 1.942 | 2.115 | 1.248 |
| | (1.005) | (1.304) | (1.144) | (1.073) | (1.305) | (1.122) | (1.277) | (1.150) |
| | 2.647 | 2.743 | 2.119 | 2.973 | 1.549 | 1.730 | 1.656 | 1.085 |
| | 0.024 | 0.024 | 0.07 | 0.024 | 0.141 | 0.132 | 0.132 | 0.279 |
| Use length AAS> 10 | 4.149*** | 3.972** | 2.779* | 1.509 | 3.427* | 2.159 | 3.939** | 2.811* |
| | (1.079) | (1.406) | (1.225) | (1.152) | (1.403) | (1.203) | (1.376) | (1.225) |
| | 3.846 | 2.824 | 2.268 | 1.309 | 2.442 | 1.795 | 2.863 | 2.295 |
| | <.001 | 0.013 | 0.032 | 0.192 | 0.03 | 0.085 | 0.013 | 0.032 |
| Time | 0.220 | 0.210 | -0.855 | -0.835 | -0.471 | 0.003 | 0.830 | 0.756 |
| | (0.511) | (0.721) | (0.564) | (0.549) | (0.681) | (0.555) | (0.694) | (0.521) |
| | 0.431 | 0.292 | -1.515 | -1.521 | -0.692 | 0.005 | 1.195 | 1.451 |
| | 0.881 | 0.881 | 0.397 | 0.397 | 0.784 | 0.996 | 0.47 | 0.397 |
| Age | -0.038 | -0.011 | -0.037 | -0.001 | -0.009 | -0.018 | -0.046 | -0.074 |
| | (0.051) | (0.065) | (0.058) | (0.054) | (0.066) | (0.057) | (0.064) | (0.058) |
| | -0.744 | -0.171 | -0.645 | -0.020 | -0.137 | -0.318 | -0.712 | -1.266 |
| | 0.984 | 0.984 | 0.984 | 0.984 | 0.984 | 0.984 | 0.984 | 0.984 |
| Observations | 305 | 305 | 305 | 305 | 305 | 305 | 305 | 305 |
| Log Likelihood | -985.282 | -1,070.670 | -1,022.598 | -1,005.761 | -1,066.807 | -1,016.968 | -1,063.062 | -1,018.173 |
| Akaike Inf. Crit. | 1,984.564 | 2,155.339 | 2,059.196 | 2,025.522 | 2,147.615 | 2,047.936 | 2,140.123 | 2,050.345 |
| Bayesian Inf. Crit. | 2,010.607 | 2,181.381 | 2,085.238 | 2,051.564 | 2,173.657 | 2,073.978 | 2,166.165 | 2,076.387 |
